# Age-specific burden of medically attended respiratory virus disease in high-income countries: a scoping review and meta-analysis

**DOI:** 10.64898/2026.06.09.26354660

**Authors:** Muskaan Gupta, Hordur Zoega, Isaac J. Stopard, Bette Liu, Kristine Macartney, James G Wood, Alexandra B Hogan

## Abstract

**Introduction:** Respiratory infections are a leading cause of morbidity. Newly available vaccines to prevent respiratory syncytial virus (RSV) disease and encouraging clinical progress on vaccines for human metapneumovirus (hMPV) and parainfluenza (PIV) could reduce the disease burden beyond existing influenza and SARS-CoV-2 immunisation programs. However, evidence on the contribution of these viruses to respiratory disease burden across the lifespan remains limited.

**Methods:** We reviewed studies from 01/2002–11/2025 reporting age-stratified, medically attended cases of influenza, and at least one of RSV, hMPV, or PIV, in high-income countries, excluding periods substantially overlapping with the COVID-19 pandemic. Using only studies that tested for all four viruses, we estimated the age-specific proportion of cases that were non-influenza (total across RSV, hMPV and PIV) compared to influenza using a mixed-effects logistic regression model.

**Results:** Following exclusions and screening, 61 studies were included in the primary analysis comprising >500,000 detections of the four viruses. We found that a substantial proportion of medically attended respiratory illness in infants and young children was due to PIV, hMPV and RSV, rather than influenza, with a non-influenza virus proportion of 90.2% (95% CI 85.9–93.2%) in young infants aged 0–6 months. The converse was true for school-aged children, with a non-influenza virus proportion of 34.8% (95% CI 26.5–44.2%) in children aged 5–18 years. In adults aged 65+ years, non-influenza causes of medically attended disease were common at 60.2% (95% CI 50.0–69.5%). Restricting to studies reporting hospitalised cases (n=19) produced broadly similar age-specific trends in relative virus burden contributions.

**Discussion:** We highlight the significant burden of medically attended illness due to PIV, hMPV and RSV across ages, particularly in infant and preschool-aged children and older adults, supporting the need for effective vaccines targeting this burden.

## Introduction

Respiratory viruses are a leading cause of hospitalisations worldwide and contribute significantly to global morbidity, mortality and healthcare costs.^1,2^ Severe acute respiratory syndrome coronavirus 2 (SARS-CoV-2), respiratory syncytial virus (RSV), influenza, human metapneumovirus (hMPV), and parainfluenza (PIV) are major contributors to this global burden.^3^ The incidence and clinical impact of these infections vary substantially with age, reflecting changes in individual-level immunity and susceptibility over time.^4,5^

Existing studies of virus burden generally focus on single virus analyses within restricted age bands (e.g., RSV in younger children or hMPV in older adults).^6,7^ However, few studies have integrated age-stratified data across multiple viruses, even in high-income settings where medically attended case data is more systematically recorded. Consequently, although the burden of specific respiratory viruses is well-described in certain populations (particularly RSV in infants and young children), comparative evidence on the burden and severity across the lifespan remains limited.^8^

Understanding the age-specific virus burden and clinical impact is especially relevant as several of these viruses are already, or could become, vaccine preventable. Identifying which viruses are most prevalent in different age groups could also inform the future implementation of combination vaccines, which target two or more pathogens in a single formulation.^9^ Combination vaccines are widely used in infant and childhood immunisation programmes (e.g. hexavalent and measles-mumps-rubella vaccines), but their use is currently more limited in older populations.^10^ Targeted combination respiratory virus vaccination could help to streamline immunisation schedules, enhance coverage against age-specific viral threats, and improve health system efficiency.^10^

In this study we undertook a scoping review and meta-analysis to quantify the age-stratified burden of RSV, hMPV, PIV and influenza disease in high-income settings, and used these data to quantify how the relative burden of these viruses changes over the life course. We summarised the current development landscape of combination vaccines for RSV, hMPV, PIV, influenza and SARS-CoV-2 to contextualise our findings.

## Methods

### Search strategy

We searched PubMed for studies dating from January 2002 up to November 2025 for English-language, human studies reporting on the number of medically confirmed positive cases for two or more of the following respiratory viruses: influenza, RSV, PIV, and hMPV. We specifically did not include SARS-CoV-2 in the search because it circulated only for a small proportion of the review time period and its stable endemic transmission pattern is not yet well characterised. Additional studies were identified by screening reference lists of relevant identified articles. The full search strategy is detailed in the Supplementary Information, with the search and screening undertaken by one individual.

Titles and abstracts were screened for relevance using EndNote 21.2, and the full texts of remaining articles were assessed against the inclusion and exclusion criteria. Studies were included if they were based on data from high-income countries as per the World Bank’s 2025-2026 classification^11^; reported on positive cases for respiratory viruses stratified by age groups; and had a minimum of one positive influenza case in at least one age group. Studies were excluded if they had a testing period that substantially overlapped (>50% of the reported testing period) with the COVID-19 pandemic (March 2020–May 2023). This restriction was applied to minimise the influence of pandemic-related disruptions on respiratory virus circulation, as data from this period are unlikely to be representative of typical viral transmission patterns and underlying burden.

### Data extraction and classification

Study details (authors, year, study design, country), study methods (inclusion criteria, age-stratification ranges, testing method, study period, viruses reported) and outcomes (study setting, age group, positive case counts by virus) were extracted and reviewed by an independent reviewer. Where data were presented in percentages of a total case count, a back-calculation was used to estimate raw case counts. Further, when data were presented only graphically, values were extracted (where feasible) using *WebPlotDigitizer.*^12^

Cases were coded by severity through full-text review to assess the influence of disease severity on estimates. A case was classified as mild if the patient was tested only in community or outpatient settings, and as severe if the patient was hospitalised due to respiratory infection, discussed further in the Supplementary Information.

To enable consistent comparison across studies, age groups were standardised into six categories: young infants (0–6 months), toddlers (6–24 months), preschoolers (2–5 years), children (5–18 years), adults (18–65 years), and older adults (65+ years). An additional “infant toddler” category was used for studies that reported cases for broader age ranges that partially overlapped the young infant and toddler groups, defined as including any portion of the first 6 months of life, and extending beyond the 6–12-month range. Where study age categories did not align with our classification, we assigned the category that most closely aligned; these assumptions are detailed in the Supplementary Information.

### Data analysis

To evaluate the relative contribution of each virus to the overall medically attended disease burden, we first restricted the data to only studies that tested for all of RSV, hMPV, PIV, and influenza. For each study and age group, we calculated the proportion of positive cases attributable to RSV, hMPV, PIV, and influenza out of the total number of positive cases across all four viruses.

We then explored the relationship between age and the proportion of cases due to non-influenza viruses (i.e. RSV, hMPV, PIV), relative to all positive cases including influenza. Influenza was used as the comparator because, among the viruses analysed, it has the most well-characterised age-specific burden. Additionally, vaccination programs for influenza are well-established, and there is scope for future additional respiratory virus vaccinations to be combined or co-administered with influenza vaccination.

We examined the relationship between age and the proportion of cases due to non-influenza viruses using a mixed-effects logistic regression model, where the binomial outcome was the number of non-influenza cases out of the total cases, indexed by age and study. Age was included as a fixed effect, with a random intercept included to capture study level differences. The model is described as

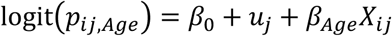

where *p* is the probability of a non-influenza case, *β*_0_ is the intercept, *u*_*j*_ is the random effect for study *j*, *β*_*Age*_ is the fixed effect on the intercept for each age group, and *X*_*ij*_represents each observation *i* within study *j*. This model allowed us to estimate the probability of a test being positive for a non-influenza virus (given that the test is positive for either non-influenza or influenza), for each age group of interest. Models were fitted using the `glmer()` function in the `lme4` package in R version 4.5.2.^13^ To estimate absolute age-specific proportions, we added the coefficient for each age-group to that for the reference group to represent the log-odds for that age group, and then transformed these estimates to the probability scale using the inverse logit function `plogis()`.

In an additional analysis, we used the same approach to explore this relationship on a per-virus basis for each of RSV, hMPV and PIV in comparison to influenza, without the restriction that studies needed to test for all four viruses of interest.

Finally, we considered whether the relative age-specific burden would change if we restricted our analysis to studies of hospitalised cases or emergency department presentations only (i.e. restricting to severe disease).

### Baseline influenza burden

To contextualise our results, we extracted 2023–2024 influenza hospitalisation data from three exemplar high-income countries where age-stratified hospitalisation rates were available (Australia, United States and United Kingdom).^14–17^ The 2023–2024 influenza season was chosen as a reference as it was the most recent year with available data.

These estimates relied on different surveillance approaches: Australian data were produced directly from coded hospital separation statistics, US data were produced via modelled extrapolation of hospital sentinel surveillance, and UK data were produced directly from sentinel hospital surveillance data in defined catchment regions. This baseline influenza hospitalisation burden is shown as a reference only and was not otherwise used in our analysis (see Supplementary Information for further details).

### Combination vaccine landscape

To further frame our findings, we reviewed the clinical development landscape for combination vaccines targeting influenza, RSV, hMPV, PIV, and SARS-CoV-2. To do this, we conducted a comprehensive search of two major clinical trial registries: ClinicalTrials.gov,^18^ and the World Health Organisation’s (WHO) International Clinical Trials Registry Platform (ICTRP).^19^ Trial characteristics were synthesised and visualised to provide an overview of the current landscape of respiratory vaccine development. Further details of the review methods are provided in the Supplementary Information.

## Results

### Search strategy and data extraction

From the virus burden search, we initially identified 14,511 articles. After duplicates were excluded, titles and abstracts were screened for 11,843 articles, and we subsequently assessed 326 studies for eligibility. A total of 265 studies were excluded, with the most common exclusion reasons being that data were not for high-income countries (n=136), data were not age-stratified (n=80), or case counts were either not reported or unable to be calculated from the data presented (n=21). We excluded a further two studies that overlapped with a dataset that was already captured in a third study in the review.^20,21^ One study was excluded because it reported on modelled data rather than raw case counts.^22^ After study exclusions (summarised in Figure 1), 61 studies were included in the analysis (Table S3), representing a total of 23 countries. Studies reported a median of four numerical age categories (range 1–9, IQR 3–6). Once consolidated into this review’s descriptive age categories, this yielded a median of three age groups per study (range 1–5, IQR 2–4).

**Figure 1.**
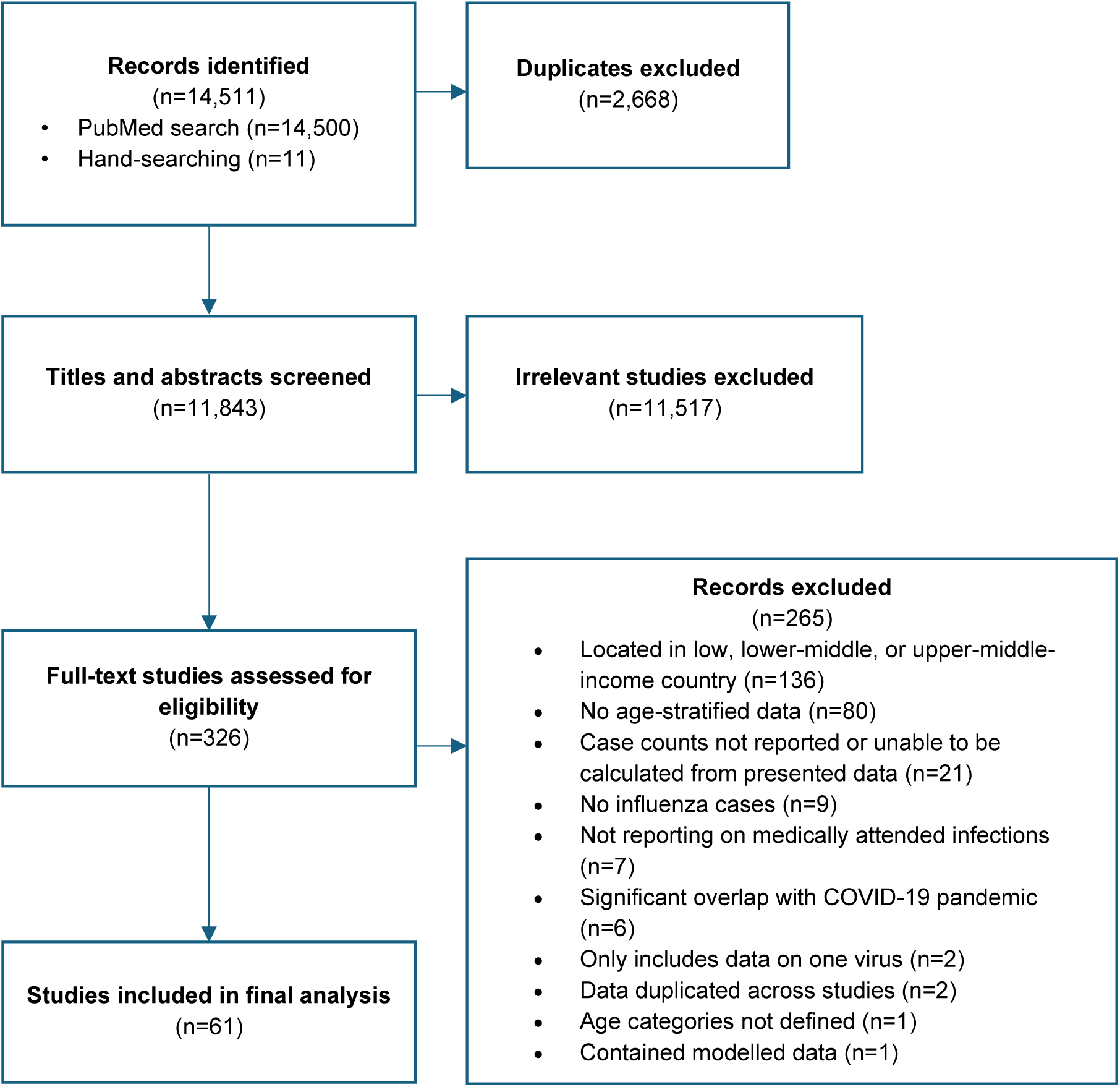
PRISMA flowchart for the virus burden literature review.

### Age-specific burden of non-influenza viruses relative to influenza

Of the 61 eligible studies, 29 tested for all four respiratory viruses of interest. We observed that the relative incidence of hMPV and PIV cases was consistent across the age spectrum, whereas the relative incidence of RSV was greatly elevated in younger age groups. Conversely, the relative burden of influenza was higher in age groups older than preschoolers, including school-aged children up to older adults (Figure 2A).

**Figure 2.**
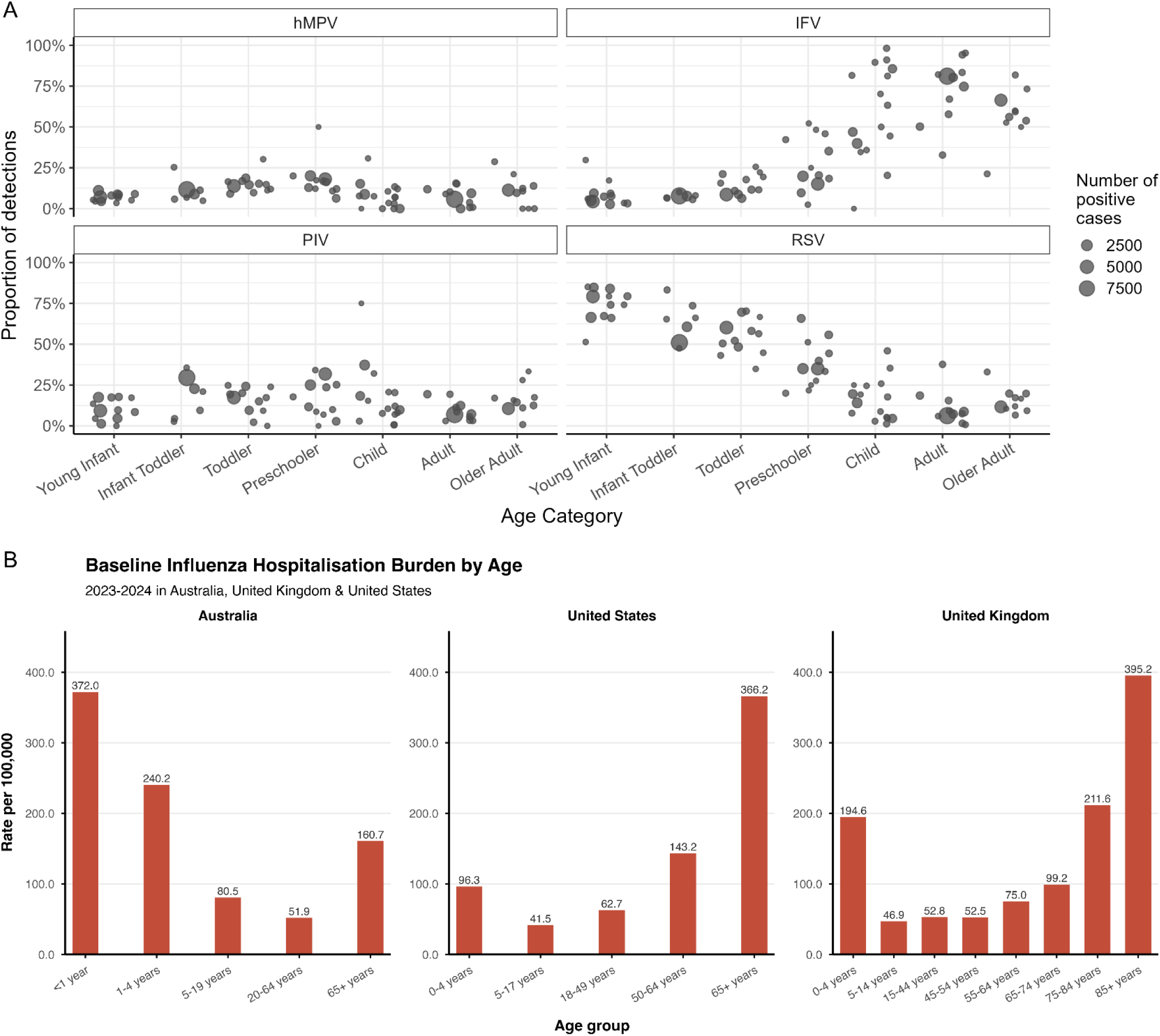
Age-specific proportional detections of hMPV, influenza, PIV and RSV, and age-specific influenza hospitalisation data, in high-income countries. A) Age-specific proportion of detections for each of the hMPV, influenza, PIV and RSV, out of the total positive detections across the four viruses, where only studies featuring testing for all four viruses are included. Age groups: young infant (0–6 months); infant toddler (partial overlap across young infant and toddler groups); toddler (6–24 months); preschooler (2–5 years); child (5–18 years); adult (18–65 years); and older adult (65+ years). B) Influenza virus hospitalisation rates in Australia, the United States, and the United Kingdom, per age group (2023–2024)^14–17^.

Absolute influenza-associated hospitalisation rates across the exemplar settings of Australia, the UK and the US demonstrated a consistent U-shaped age pattern, with the highest rates observed in children younger than 5 years, and adults aged 65 years and older. The risk of influenza hospitalisation in individuals aged 5 years to 65 years was consistently low across settings. The magnitude of the age gradient in young children and older adults varied between settings; in particular, Australia showed the highest hospitalisation rates in infants (Figure 2B).

We applied a mixed-effects regression model to quantify the relative age-specific proportion of positive cases not attributable to influenza. We found that relative to young infants, children aged 5–18 years and adults aged 18–64 years had the lowest odds of a non-influenza case, with odds ratios of 0.06 (95% CI 0.05–0.07) and 0.08 (95% CI 0.07–0.1) respectively, rising to 0.16 (95% CI 0.13–0.21) in older adults (aged >65 years) (Figure 3A).

**Figure 3.**
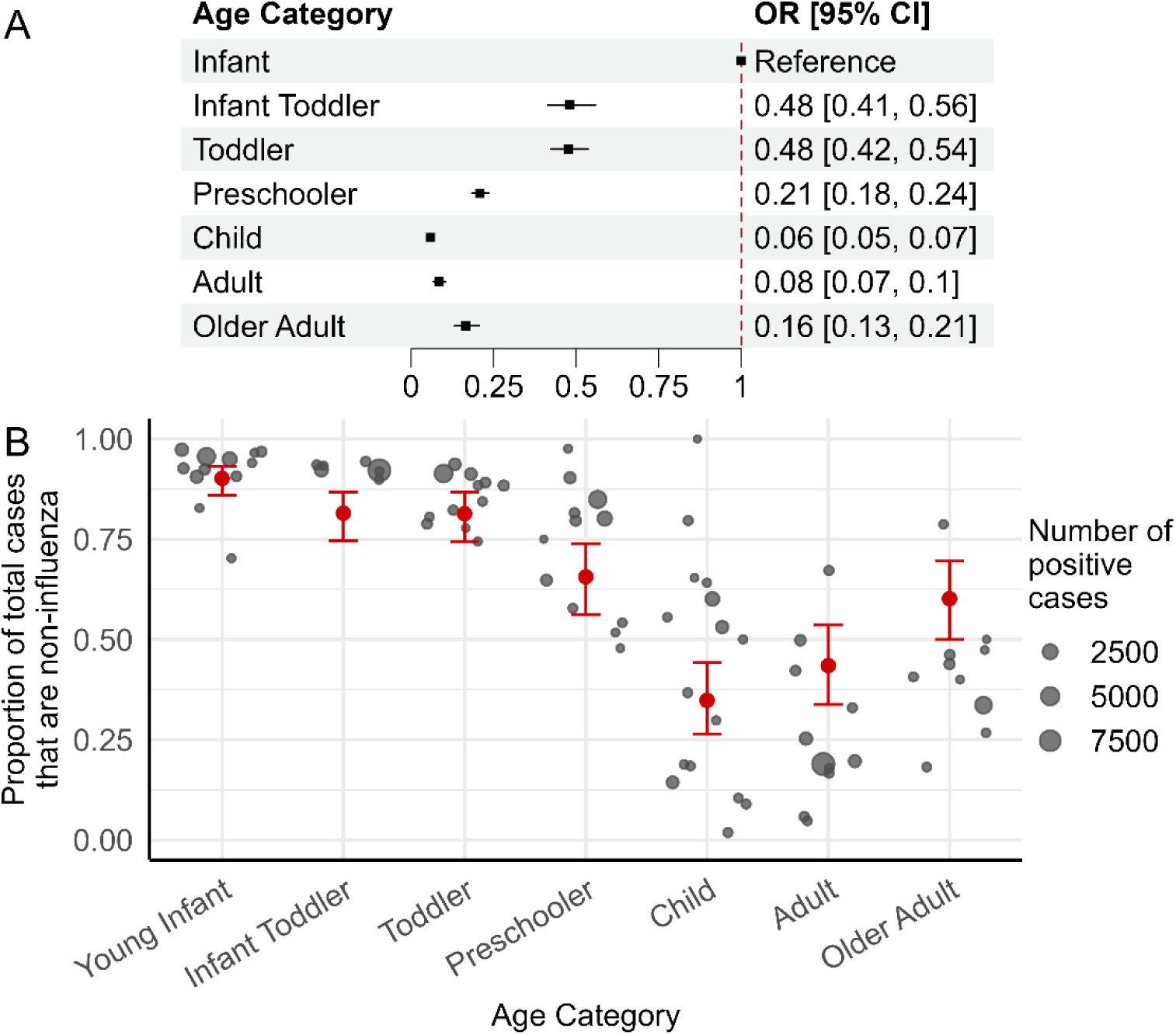
Age-specific burden of non-influenza (hMPV, PIV and RSV) relative to influenza virus. A) Forest plot of model results illustrating odds ratios for the proportion of cases that are non-influenza for each age group, relative to the young infant reference group. B) Absolute proportion of detections that are non-influenza (PIV + hMPV + RSV), relative to total burden (including influenza). Age groups: young infant (0–6 months); infant toddler (partial overlap across young infant and toddler groups); toddler (6–24 months); preschooler (2–5 years); child (5–18 years); adult (18–65 years); and older adult (65+ years).

In absolute terms, we found that the modelled proportion of cases not attributable to influenza was largest in young infants (90.2%, 95% CI 85.9–93.2%), with a slightly lower proportion of non-influenza cases in toddlers (81.4%, 95% CI 74.4–86.7%), falling further in preschool children (65.6%, 95% CI 56.2–73.9%). In school-aged children and adults, the proportion of cases attributable to the three other respiratory viruses was lower than that attributable to influenza, with non-influenza proportions of 34.8% (95% CI 26.5–44.2%) in school-aged children and 43.5% (95% CI 33.8–53.6%) in working-age adults. In adults aged 65+ years, non-influenza causes of medically attended disease were more common than influenza-attributable disease, with 60.2% (95% CI 50.0–69.5%) of cases attributable to the three non-influenza pathogens (Figure 3B). Absolute proportion results are summarised in Figure 5A.

In some age groups, the modelled estimate fell outside the reported data (e.g. older adults), due to inclusion of a study-specific random effect in the model. Direct comparisons between observed study-level values and the modelled estimate are shown in Figure S4.

### Age-specific burden of each of RSV, hMPV and PIV relative to influenza

We fitted the same model to explore the relationship between virus burden and age on a per-virus basis for each of RSV, hMPV and PIV in comparison to influenza, without the restriction that studies needed to test for all four viruses of interest (i.e. capturing all 61 studies) (Figure 4). We found that for RSV, relative to young infants, all other age groups had low odds of a respiratory case being attributable to RSV, with the lowest in children and adults (odds ratio 0.01 for both groups, 95% CI 0.01–0.01 for both), and an odds ratio of 0.02 in older adults (95% CI 0.02–0.02) (Figure 4).

**Figure 4:**
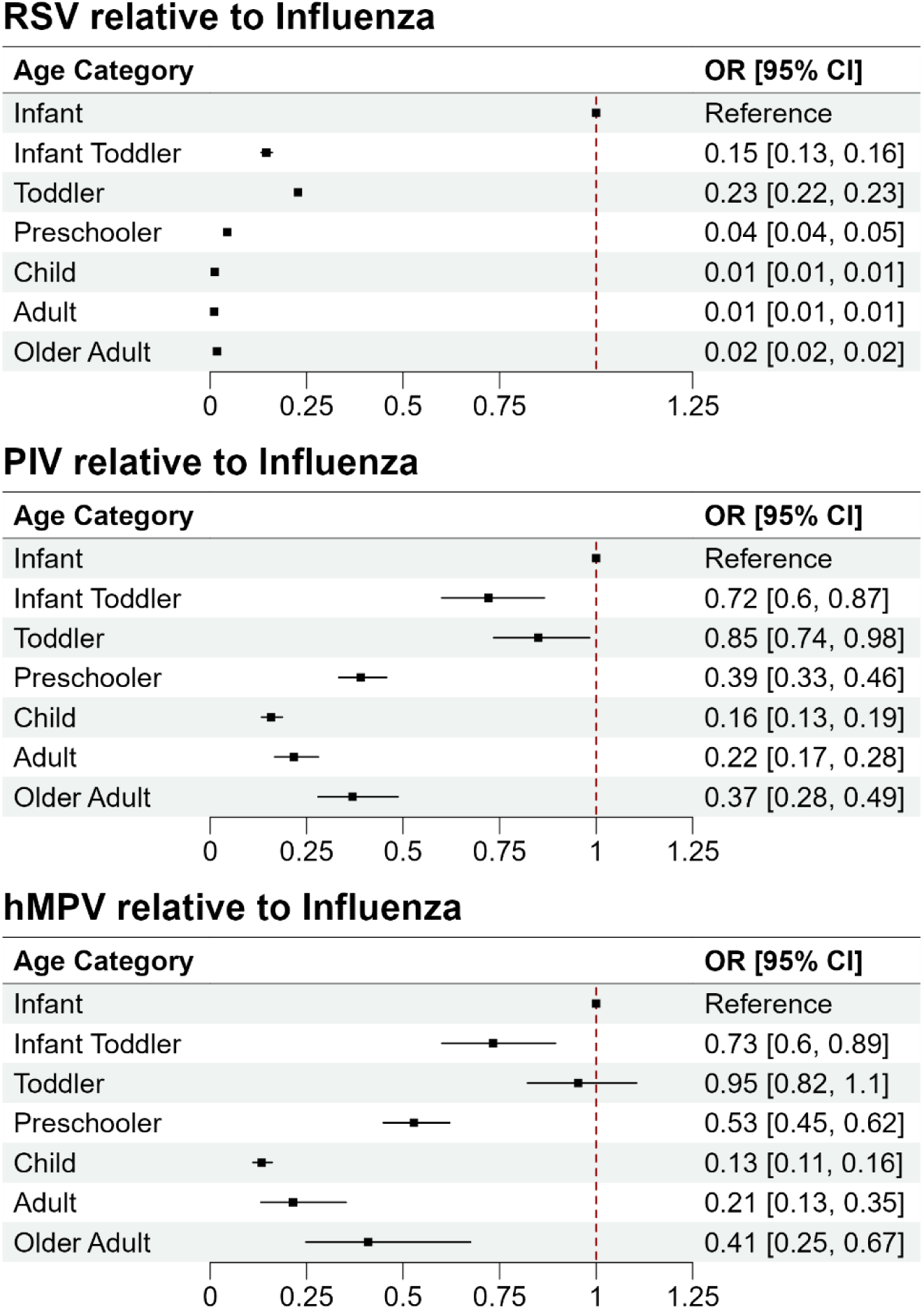
Age-specific burden of non-influenza (hMPV, PIV and RSV) relative to influenza virus, with all reviewed studies (. ***n*** = **61** ). Forest plot of model results illustrating odds ratios for the proportion of cases that are non-influenza for each age group, relative to the young infant reference group. Age groups: young infant (0–6 months); infant toddler (partial overlap across young infant and toddler groups); toddler (6–24 months); preschooler (2–5 years); child (5–18 years); adult (18–65 years); and older adult (65+ years).

**Figure 5:**
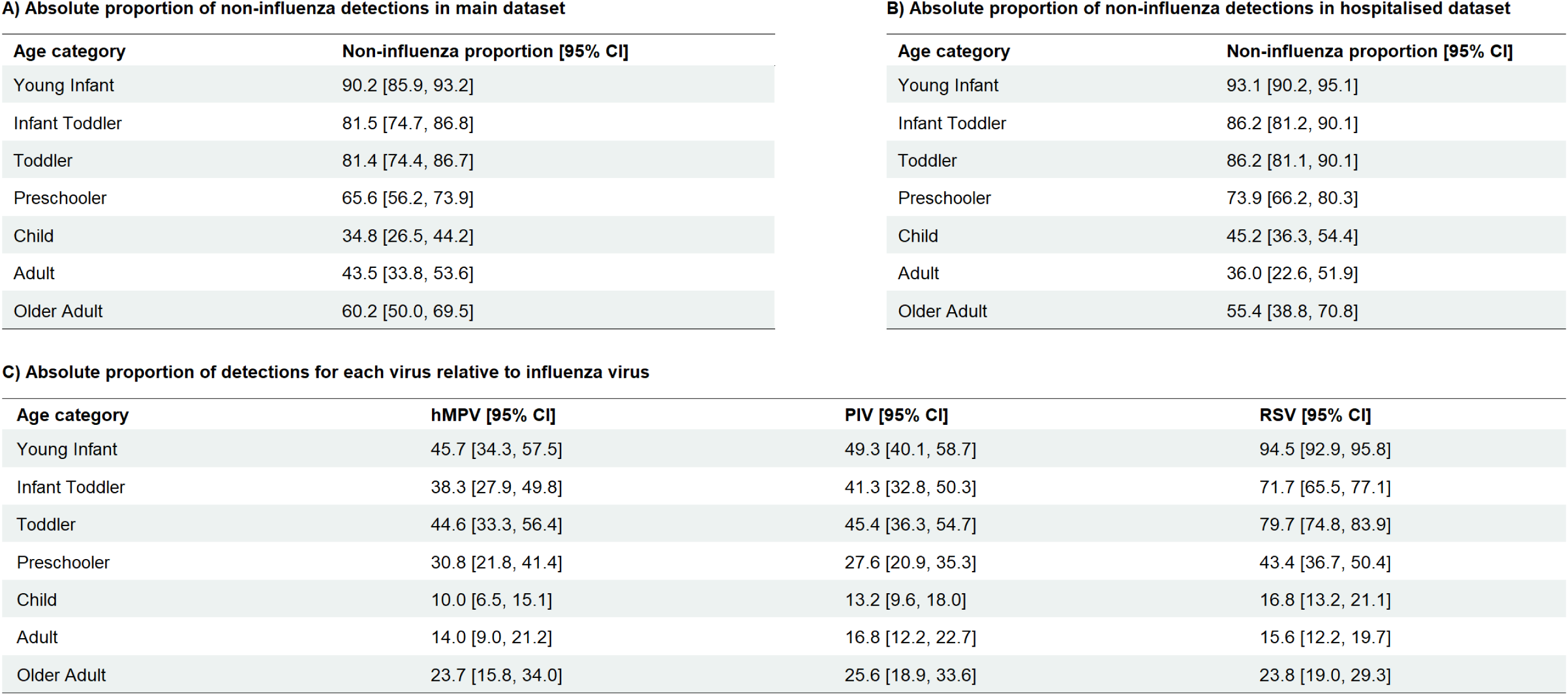
Age-specific virus burden of hMPV, PIV, RSV and influenza virus. A) Age-specific burden of non-influenza (hMPV, PIV and RSV) relative to influenza virus, with all reviewed studies (*n* = 61). Corresponds to the model fit (red) in Figure 3B in and Figure S2 (supplemental information). B) Age-specific burden of non-influenza (hMPV, PIV and RSV) relative to influenza virus, restricted to hospitalised dataset (*n* = 19). Corresponds to the model fit (blue) in Figure S2 (supplemental information).C) Age-specific proportion of detections for each of hMPV, PIV and RSV, relative to influenza virus (*n* = 61). Corresponds to the model fit (red) in Figure S3 (supplemental information). Age groups: young infant (0–6 months); infant toddler (partial overlap across young infant and toddler groups); toddler (6–24 months); preschooler (2–5 years); child (5–18 years); adult (18–65 years); and older adult (65+ years).

When examining the model fitted on a per-virus basis, we found that PIV and hMPV exhibited similar age-specific trends relative to young infants. We estimated that the odds of a case being PIV and hMPV was similar in toddlers to in young infants (odds ratio 0.85 (95% CI 0.74–0.98) and 0.95 (95% CI 0.82–1.1) for PIV and hMPV respectively). In children and adults, influenza was much more likely compared to in infants (Figure 4). For PIV, we estimated an odds ratio of 0.16 in children relative to young infants (95% CI 0.13–0.19), and of 0.22 in adults (95% CI 0.17–0.28). For hMPV, we estimated odds ratios of 0.13 (95% CI 0.11–0.16) and 0.21 (95% CI 0.13–0.35) in children and adults respectively. In absolute terms, we found similar age-specific burdens for both hMPV and PIV in comparison to influenza (Figure 5, Figure S3). In adults and older adults, the relative ratios of cases attributed to RSV, hMPV and PIV compared to influenza were all similar (Figure 5, Figure S3).

### Age-specific burden of non-influenza viruses relative to influenza in hospitalised cases only

Restricting our dataset to only include hospitalised cases (excluding emergency department attendances discharged without admission, but retaining those admitted or with unknown admission status) resulted in 19 studies for analysis. The model fit to these studies only (compared to the full dataset) exhibited a similar directional trend, with some differences in point estimates and largely overlapping confidence intervals. The largest difference was in preschoolers and children, where the proportion of non-influenza virus detections was higher for the hospitalised dataset at 73.9% (95% CI 66.2–80.3) for preschoolers and 45.2% (95% CI 36.3–54.4) for children, compared to 65.6% (95% CI 56.2–73.9) for preschoolers and 34.8% (95% CI 26.5–44.2) for children in the full dataset (Figure 5B, Figure S2)

### Combination vaccine landscape

A total of 22 vaccines were identified in active clinical development. Of these, 18 vaccine candidates targeted two pathogens, with seven combining RSV and hMPV, six combining influenza and SARS-CoV-2, two combining hMPV and PIV-3, and one combining RSV and influenza. The remaining four candidates targeted three pathogens, with three including RSV, hMPV and PIV-3 and one including influenza, SARS-CoV-2 and RSV (Figure S5).

Most vaccine candidates were in early stage of development. RSV–hMPV and influenza–SARS-CoV-2 combinations were the only formulations having progressed to Phase 2 clinical trials, with two influenza–SARS-CoV-2 vaccines advanced to Phase 3. A range of platforms were represented, with nucleic acid or subunit platforms most commonly evaluated. Thus far, relatively few combinations have been tested in paediatric groups, with most trials enrolling adults only.

## Discussion

In this paper we reviewed studies of disease burden in high-income countries involving laboratory testing of influenza and at least one of RSV, PIV or hMPV. Our analysis focused on these four pathogens as they collectively represent a large burden of clinically significant viral respiratory disease globally and are potential targets for future combination vaccines and other new vaccine products. Using data from these studies, we estimated the relative age-specific burden of medically attended cases attributable to these four diseases in high-income countries. Our findings support the established dominance of RSV in infants and toddlers, and influenza as a prominent respiratory pathogen from school age through to older adulthood.

Influenza is traditionally regarded as the primary source of severe viral respiratory morbidity in older adults, with RSV more recently highlighted as a major source of disease in this population.^23,24^ However, the prominence of PIV and hMPV has not been recognised to the same extent.^25–27^ Our analysis demonstrates that PIV and hMPV contribute to substantial respiratory disease in adult and older adult age groups, with a medically attended burden similar to that for RSV. This finding underscores the importance of non-influenza respiratory viruses beyond early childhood, and suggests that PIV and hMPV represent important targets for respiratory virus vaccines, particularly in light of the recent approval for RSV vaccines for older adults.^28^

Because the catchment population size for each study was generally unknown, we presented influenza disease burden as relative to the total cases or influenza cases. To provide additional context for our results, we illustrated the age-specific baseline influenza hospitalisation in Australia, the United Kingdom and the United States. All settings were characterised by a U-shaped pattern of influenza hospitalisation risk by age, with the highest risk in young children and older adults. While there could be variation in risk of influenza disease in different populations, these differences between countries likely also reflect geographical variation in testing practices, influenza-attributable coding differences, and seasonal strain dominance.

Our statistical analysis pointed to a similar medically attended burden of hMPV and PIV in infants and young children compared to that for influenza; the high influenza hospitalisation rate in this age group suggests that the absolute burden of these pathogens could be substantial and that an effective combined hMPV-PIV vaccine may potentially avert a considerable proportion of respiratory disease, as would a paediatric RSV vaccine. However, despite the substantial burden of hMPV, PIV and RSV in young children, clinical evaluation of combination vaccines for these pathogens in young children is limited). This gap likely reflects challenges in conducting paediatric clinical trials, including in relation to ethical, recruitment, and scientific challenges (such as population heterogeneity).^29^ The scarcity of trials in paediatric populations highlights a disconnect between current development pathways and the populations that stand to benefit most from effective prevention strategies. In addition, effective vaccination in early childhood and school-aged groups may confer indirect benefits through reduced transmission in the community, increasing the population-level effectiveness of such strategies.

Our study has important strengths. Presenting age-specific burden across the full lifespan, and compared to influenza, supports interpretability and helps clarify the public health benefit of future prevention efforts. To our knowledge, few studies have comprehensively synthesised the age-stratified burden for hMPV and PIV across the life course. Grouping pathogens together in ways that represent plausible combination formulations, complemented by an overview of the respiratory vaccine development pipeline, offers a direct indication of the potential added value of pipeline vaccine products in different age groups of interest.

Our study has several limitations. First, our included studies featured a range of study designs over a period of about 20 years, with some variation in case definitions, testing procedures and outcomes. To partially address consistency of severity between studies, we conducted additional sensitivity analysis where we restricted to studies reporting hospitalised cases.

Second, we note potential differences between country settings in terms of respiratory virus epidemiology, which can be driven by setting-specific factors such as seasonality, population structure and access to care, which may limit generalisation. We restricted our search criteria to studies based on data for high-income settings only, to partially address this limitation. Third, many studies only captured data for a subset of the age-range (e.g. either reporting cases in infants, children, older adults) with only a small number of studies reporting cases in both younger children and adults. We assigned our broad age categorisation to help aid interpretability and applicability of our statistical model outcomes but note lack of perfect overlap with many of the studies and that other acceptable age categorisations were possible.

A further limitation is the exclusion of SARS-CoV-2 from the analysis, despite SARS-CoV-2 now being a major endemic respiratory virus globally. This reflects the fact that most studies captured in our search were conducted prior to the COVID-19 pandemic; because our methodology required the same viruses being captured across the full analysis window, including SARS-CoV-2 would have restricted the analysis to the post-pandemic period only (a much smaller evidence base). Further, the SARS-CoV-2 pandemic and non-pharmaceutical interventions resulted in a temporary interruption to the transmission of other pathogens in many settings, meaning that studies from this period are unlikely to reflect typical transmission dynamics.^30^ As additional data accumulates over time and the transmission dynamics of endemic viruses stabilise, future analyses could incorporate SARS-CoV-2 alongside other viruses.

Gaps remain in our understanding of the burden of respiratory virus disease across the lifespan. In our study, we synthesised available evidence on four key respiratory pathogens within high-burden settings. Future research could extend this approach to lower-income strata to provide a more globally representative picture of medically attended respiratory disease burden. Findings from our study can be used to inform priority pathogens and age-targets for future vaccine development, including for combination vaccines which offer the potential for simplifying immunisation schedules and improving uptake, and health system efficiencies associated with fewer clinic visits.^10,31^ These synthesised burden estimates can also support epidemiological and modelling analyses aimed at estimating the potential population-level benefit of these future products.

## Data availability

The data and code required to reproduce the results in this manuscript in full are available on GitHub at www.github.com/abhogan/virus_burden.

## Supplementary Information

### A Respiratory Virus Burden Analysis

#### A.1 PubMed Search Strategy

**Table S1.**
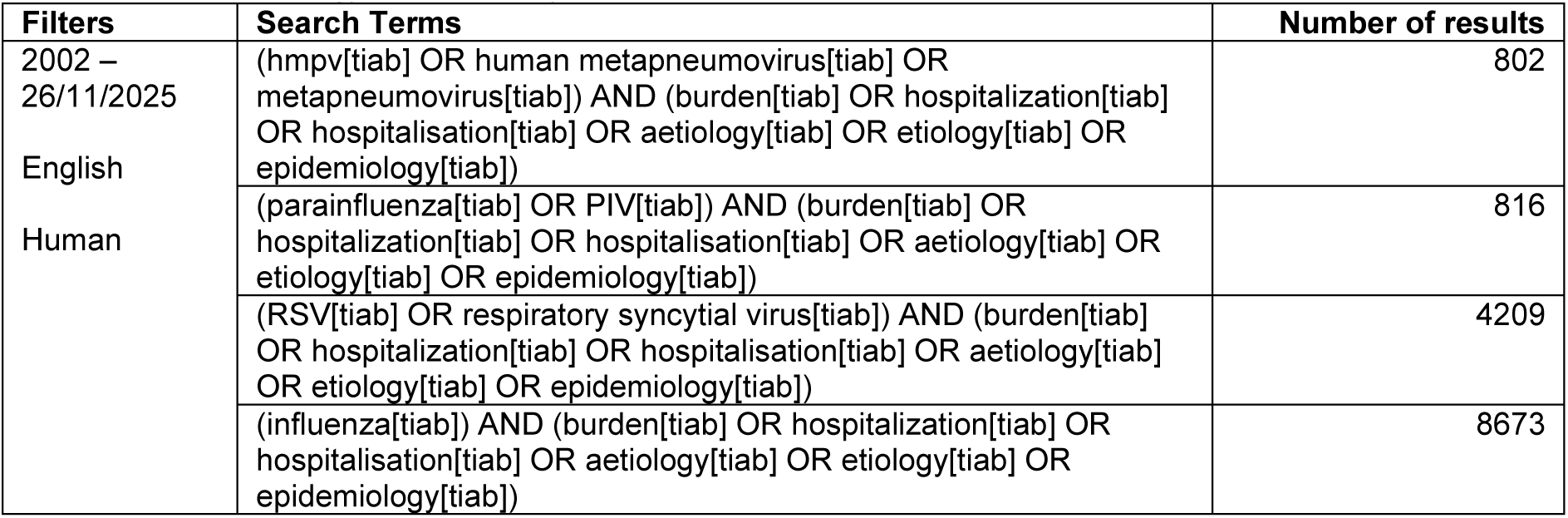
Search strategy for respiratory virus burden review.

#### A.2 Coding of Age Group

For each study and age group in which cases were reported, we coded the age group as one of seven categories: Young Infant, Infant Toddler, Toddler, Preschooler, Child, Adult, or Older Adult. Table S2 describes age ranges corresponding to each category. Where an age range captured two categories, we allocated the category that most closely aligned. For instances where age ranges captured three or more categories, we excluded that entry from the analysis: this was any age range broader than six months to four years.

**Table S2:** Description of age coding of studies in review. Y: year.

| Age Coding | Explanation |
| --- | --- |
| Young Infant | Any age group up to the first 12 months that includes the first 3 months. This included the age ranges of 0–6 months and 0–12 months. |
| Infant Toddler | Partial overlap across Young Infant and Toddler groups, where any of the first 6 months are included, as well as broader than the 6–12-month range. This includes the age ranges of 4m–12m, 0–18m, 0–3Y. |
| Toddler | Any age group in 6–24m. The age groups of 6–36m was allocated to Toddler where relevant. |
| Preschooler | Any age group in 24–60m. The age groups of 1–3Y and 1–4Y were allocated as Preschooler where relevant. |
| Child | Any age group in 5–18Y. |
| Adult | Any age group in 18–65Y. |
| Older Adult | Any age group in 65Y+. The age group of 50Y+ was allocated to Older Adult where relevant. |

#### A.3 Coding of Severity

All studies included in the review were classified into a severity category based on a full-text review (Table S3). Severity coding aimed to differentiate the severity of respiratory illness between study cohorts by separating cohorts admitted to hospital due to respiratory illness and those only treated in the outpatient/community setting. Hospitalised patients were defined in the analysis as all cohorts presenting to emergency department (excluding those clearly not admitted, but retaining those admitted from ED or with unknown admission status), admitted to hospital and/or admitted to ICU (“HOS-RI”, “ICU-RI”, “ED-RI”, “ED-RI-ADM”). Further information on individual study coding is available in the accompanying dataset.

**Table S3:** Description of severity coding of studies in review.

| Severity Coding | Explanation |
| --- | --- |
| MIX | Multiple sites used in study (e.g. hospitalisation, ED, outpatient etc.) |
| OP | Outpatient / primary care, not further specified |
| OP-SYM | Outpatient / primary care, respiratory illness symptomatic |
| OP-SURV | Outpatient / primary care, part of surveillance program |
| ED-RI | Presented to ED for RI symptoms, admission status not available |
| ED-RI-ADM | Presented to ED, for respiratory illness, then admitted to hospital |
| ED-RI-NOADM | Presented to ED, for respiratory illness, NOT admitted to hospital |
| HOS-RI | Hospitalised, for respiratory illness |
| ICU-RI | Admitted to ICU for respiratory illness |

#### A.4 Baseline Influenza Burden

The reference baseline influenza burden in high-income countries in 2023–2024 was produced using data from three countries where hospitalisation data was accessible: Australia, the United Kingdom and the United States. Age-stratified hospitalisation rates from these countries were extracted using the methods below and summarised by country in Figure 2B.

##### Australia

Australian baseline influenza burden (July 2023 to June 2024) was compiled using hospital diagnosis codes extracted from the Australian Institute of Health and Welfare (AIHW) separation statistics. Rates were calculated using population data from the Australian Bureau of Statistics (ABS).^64,69^ International Classification of Diseases – 10^th^ Revision – Australian Modification 12^th^ Edition (ICD-10-AM) primary diagnosis codes (J09, J10.0, J10.1, J10.8) were used to extract the number of influenza virus hospitalisations from the AIHW separation data.^67,68^ Influenza virus hospitalisation rates (per 100,000) were then calculated using population data from Q1 2024 to provide an indication of the age-stratified burden of influenza virus.^64^

ABS population data estimates were provided in single-year age increments for all persons in Australia (i.e. *0–1 years, 1–2 years* etc.).^64^ AIHW separation data was provided in 5-year increments, except for *under 5 years* and *over 85 years,* which were split into *age under 1* and *age 1–4 years*, and *age over 85 years*, respectively. ABS population data was aggregated to match the age groupings of the AIHW separation data.

###### United Kingdom

For the United Kingdom, we used UK Health Security Agency weekly influenza hospital admission surveillance data for England, restricted to all sexes and the hospital admission rate metric.^66^ We defined the analysis window as 1 July 2023 to 30 June 2024 and summed weekly age-specific admission rates across the 52-week period to derive season-level cumulative rates per 100,000 population. UK rates were presented using the native reported age groups: 0*–*4, 5*–*14, 15*–*44, 45*–*54, 55*–*64, 65*–*74, 75*–*84, and 85+ years.

###### United States

For the United States, we used the US CDC influenza disease burden modelled estimates dataset and restricted analyses to the 2023*–*2024 influenza season.^65^ We extracted age-specific values for hospitalisation rate. Rates were presented using the native CDC age groups: 0*–*4, 5*–*17, 18*–*49, 50*–*64, and 65+ years.

#### A.5 Study List

**Table S4.** Overview of included studies in respiratory virus burden review. IFV: influenza; ARTI: acute respiratory tract infection; PCR: polymerase chain reaction; ARI: acute respiratory infection; ILI: influenza-like illness.

| Study ID | Author | Year | Study Design | Country | Testing Period | Testing Method | Inclusion Criteria | Viruses reported |  |  |  |
| --- | --- | --- | --- | --- | --- | --- | --- | --- | --- | --- | --- |
|  |  |  |  |  |  |  |  | RSV | PIV | hMPV | IFV |
| S01 | Sarna et al. | 2024 | Retrospective | Australia | Jan 2010-Mar 2022 | PCR | Hospital admission with lab-confirmed viral infection | ✓ | ✓ | ✓ | ✓ |
| S02 | Minney-Smith et al. | 2019 | Retrospective | Australia | Jan 2012-Dec 2015 | PCR | Patients admitted to hospital with respiratory illness | ✓ | ✓ | ✓ | ✓ |
| S03 | Amer et al. | 2015 | Cross-sectional | Saudi Arabia | Feb 2008- Mar 2009 | PCR | Patients hospitalised with ARTI symptoms | ✓ | ✓ | ✓ | ✓ |
| S04 | Widmer et al. | 2012 | Prospective | United States | 2006/07-2008/09 | PCR | Hospitalised with respiratory symptoms or non-localising fever | ✓ |  | ✓ | ✓ |
| S05 | Choi et al. | 2018 | Retrospective | South Korea | Jan 2013-Dec 2015 | PCR | Hospitalised with ARI | ✓ | ✓ | ✓ | ✓ |
| S06 | Shinkai et al. | 2024 | Prospective | Japan | Sept 2018 -Oct 2019 | PCR | Hospitalised with ARTI | ✓ |  | ✓ | ✓ |
| S08 | Nolen et al. | 2020 | Prospective | United States | Nov 2016- Oct 2018 | PCR | Hospitalised with ARI | ✓ |  |  | ✓ |
| S09 | Wolf et al. | 2006 | Prospective | Israel | Nov 2001-Oct 2002 | PCR | Hospitalised with LRI | ✓ |  | ✓ | ✓ |
| S10 | Sominina et al. | 2021 | Prospective | Russia | 2018-2019 | PCR | Patients with ILI requiring hospitalisation | ✓ | ✓ |  | ✓ |
| S11 | Jennings et al. | 2004 | Prospective | New Zealand | Jul-Nov 2001 | PCR | Patients with ARI | ✓ | ✓ | ✓ | ✓ |
| S12 | Arashiro et al. | 2024 | Prospective | Japan | Apr 2011-Jul 2022 | Not specified | Patients with confirmed RSV or IFV | ✓ |  |  | ✓ |
| S13 | Davis et al. | 2022 | Prospective | New Zealand | 2012-2016 | PCR | Hospitalised patients with ARI | ✓ |  |  | ✓ |
| S14 | Symes et al. | 2025 | Prospective | England | 2023-2024 | PCR | Symptomatic ARI patients admitted to hospital | ✓ |  |  | ✓ |
| S15 | Ajayi-obe et al. | 2007 | Prospective | England | Oct 2002-Nov 2003 | PCR | Patients presenting to hospital with ILI | ✓ |  |  | ✓ |
| S16 | Buchan et al. | 2019 | Retrospective | Canada | May 2009-May 2014 | PCR, culture, immunofluorescence | Hospitalised children positive for RSV or IFV | ✓ |  |  | ✓ |
| S17 | Chorazka et al. | 2021 | Retrospective | Switzerland | 2017/18-2018/19 | PCR | Lab confirmed IFV or RSV infection admitted to hospital | ✓ |  |  | ✓ |
| S18 | Trenholme et al. | 2017 | Prospective | New Zealand | Aug 2009- Jul 2011 | PCR | All admissions to hospital screened for LRI | ✓ | ✓ | ✓ | ✓ |
| S19 | Chan et al. | 2011 | Retrospective | United States | Oct 2009-Dec 2009 | PCR | Patients hospitalised with positive NPA swab | ✓ | ✓ | ✓ | ✓ |
| S20 | Ackerson et al. | 2019 | Retrospective | United States | Jan 2011- Jun 2015 | PCR | Patients hospitalised with RSV or IFV | ✓ |  |  | ✓ |
| S22 | Al-Romaihi et al. | 2019 | Retrospective | Saudi Arabia | Jan 2012-Dec 2017 | PCR | Patients presenting with ILI | ✓ | ✓ | ✓ | ✓ |
| S23 | Al-Shehri et al. | 2005 | Case-control | Saudi Arabia | Sept 1997-Oct 2001 | ELISA, immunofluorescence assay | Patients suffering from bronchiolitis admitted to hospital | ✓ | ✓ |  | ✓ |
| S24 | Almehmadi et al. | 2025 | Cross-sectional | Saudi Arabia | Jan 2023-Dec 2024 | PCR | Patients with suspected RTI | ✓ |  |  | ✓ |
| S25 | Aminisani et al. | 2025 | Prospective | New Zealand | 2012-2023 | PCR | Patients with ARI |  |  | ✓ | ✓ |
| S26 | Balas et al. | 2025 | Retrospective | Poland | 2023-2025 | PCR | Patients diagnosed with respiratory infections caused by RSV or IFV | ✓ |  |  | ✓ |
| S27 | Begley et al. | 2024 | Prospective | United States | 2017-Mar 2020 | PCR | Patients with medically attended ARI | ✓ |  |  | ✓ |
| S28 | Begley et al. | 2023 | Prospective | United States | Sept 2016-May 2019 | PCR | Patients hospitalised with ARI | ✓ |  |  | ✓ |
| S29 | Boas et al. | 2025 |  | Norway | 2017-2020, 2023-2024 | PCR | Patients hospitalised with confirmed RSV or IFV | ✓ |  |  | ✓ |
| S30 | Bosis et al. | 2008 | Prospective | Italy | Oct 2005- Mar 2006 | PCR | Children hospitalised with wheezing | ✓ |  | ✓ | ✓ |
| S31 | Bourgeois et al. | 2012 | Prospective | United States | 2003/04-2004/05 | PCR | Patients presenting to ED with ARI | ✓ |  |  | ✓ |
| S32 | Burrell et al. | 2025 | Retrospective | Australia | Jan 2014-Dec 2019 | PCR | Patients presenting to hospital with AR symptoms | ✓ | ✓ | ✓ | ✓ |
| S33 | De Luca et al. | 2022 | Retrospective | Italy | Sept 2015-Mar 2019 | PCR | Patients with confirmed viral RTI requiring PICU admission | ✓ | ✓ | ✓ | ✓ |
| S34 | Fagbo et al. | 2016 | Retrospective | Italy | Jan 2012-Dec 2013 | PCR | Patients presenting with ARI symptoms | ✓ | ✓ | ✓ | ✓ |
| S35 | Garcia-Garcia et al. | 2012 | Prospective | Spain | Sept 2004-July 2010 | PCR | Patients hospitalised with CAP | ✓ | ✓ | ✓ | ✓ |
| S36 | Gooskens et al. | 2014 | Retrospective | The Netherlands | 2006-07 | PCR | Patients with ARTI presenting to ED | ✓ |  |  | ✓ |
| S37 | Hombrouck et al. | 2012 | Prospective | Belgium | 2009-10 | PCR | Patients presenting with ILI to GP | ✓ | ✓ | ✓ | ✓ |
| S38 | Kim et al. | 2013 | Retrospective | South Korea | Dec 2006-Nov 2010 | PCR | Patients with respiratory illness | ✓ | ✓ | ✓ | ✓ |
| S39 | Kusel et al. | 2006 | Longitudinal Prospective | Australia | July 1996- July 1999 | PCR | Patients hospitalised for ARI | ✓ | ✓ |  | ✓ |
| S40 | Lauderdale et al. | 2005 | Prospective | Taiwan | Dec 2001-Apr 2002 | Serologic assays | Patients admitted with diagnosis of LRI | ✓ | ✓ |  | ✓ |
| S41 | Lee et al. | 2020 | Retrospective | South Korea | Jan 2010- Dec 2015 | PCR | Patients hospitalised with CAP | ✓ | ✓ | ✓ | ✓ |
| S42 | Mansbach et al. | 2008 | Prospective | United States | Dec 2005- Mar 2006 | PCR | Patients with bronchiolitis | ✓ |  | ✓ | ✓ |
| S44 | Puzelli et al. | 2009 | Prospective | Italy | 2004-07 | PCR | Patients presenting with ILI to GP | ✓ | ✓ |  | ✓ |
| S45 | Richter et al. | 2016 | Prospective | Cyprus | Nov 2010- Oct 2013 | PCR | Children hospitalised with ARI | ✓ | ✓ | ✓ | ✓ |
| S46 | Russell et al. | 2018 | Retrospective | United States | Oct 2011-Aug 2014 | PCR | Patients with ILI symptoms presenting to outpatient clinics | ✓ | ✓ | ✓ | ✓ |
| S47 | Tramuto et al. | 2015 | Cross-sectional, retrospective | Italy | Jul 2009-Dec 2012 | PCR | Patients admitted to ICU with ILI/ARTI | ✓ | ✓ |  | ✓ |
| S48 | Turunen et al. | 2015 | Retrospective | Finland | Jun 2007- Mar 2009 | PCR | Patients with first wheezing episode presenting to hospital | ✓ | ✓ |  | ✓ |
| S49 | Varghese et al. | 2018 | Prospective | Australia | Jan 2010- Dec 2013 |  | Patients presenting with ILI to GP | ✓ | ✓ | ✓ | ✓ |
| S50 | Vila et al. | 2022 | Retrospective | Spain | Oct 2012-Dec 2020 | Direct immunofluorescence assay, PCR | Patients with lab-confirmed viral LRTI requiring hospitalisation | ✓ | ✓ | ✓ | ✓ |
| S51 | Visseaux et al. | 2017 | Retrospective | France | May 2011- Apr 2016 | PCR | Patients tested for respiratory viruses at hospital | ✓ | ✓ | ✓ | ✓ |
| S52 | Walker et al. | 2014 | Retrospective | United States | Apr 2009- Mar 2010 | PCR | Patients with positive respiratory viral panel hospitalised | ✓ |  | ✓ | ✓ |
| S53 | Windsor et al. | 2022 | Retrospective | United States | Dec 2018-Dec 2019 | PCR | Patients presenting with ARI symptoms to ED | ✓ | ✓ | ✓ | ✓ |
| S54 | Yen et al. | 2019 | Retrospective | Taiwan | Dec 2016- Dec 2018 | PCR | Febrile hospitalised patients with RVP test | ✓ | ✓ | ✓ | ✓ |
| S55 | Zimmerman et al. | 2015 | Prospective | United States | Dec 2012- Mar 2013 | PCR | Patients presenting to primary care centre for treatment of URI | ✓ |  |  | ✓ |
| S56 | Zimmerman et al. | 2014 | Prospective | United States | Jan-Apr 2012 | PCR | Patients presenting to primary care centre for treatment of URI | ✓ |  | ✓ | ✓ |
| S57 | Loubet et al. | 2025 | Prospective | France | 2012/13- 2021/2022 | PCR | Hospitalised with community acquired ILI | ✓ |  | ✓ | ✓ |
| S58 | Anton et al. | 2016 | Prospective | Spain | 2006/07- 2011/12 | PCR | Outpatients with ILI | ✓ | ✓ | ✓ | ✓ |
| S59 | Bosch Castells et al. | 2024 | Prospective | Spain | 2011/12- 2018/19 | PCR | Patients hospitalised with respiratory illness | ✓ | ✓ | ✓ | ✓ |
| S60 | Chen et al. | 2014 | Retrospective | Taiwan | Jan 2009-Mar 2011 | PCR | Patients hospitalised with acute bronchiolitis | ✓ | ✓ | ✓ | ✓ |
| S61 | Cohen et al. | 2024 | Retrospective | Israel | Jan 2011-Jul 2021 | PCR | Patients admitted to PICU with laboratory-confirmed respiratory viral infection | ✓ | ✓ | ✓ | ✓ |
| S62 | De Conto et al. | 2019 | Retrospective | Italy | Oct 2012-Sept 2015 | Indirect immunofluorescence assay | Patients with respiratory illness and fever | ✓ | ✓ | ✓ | ✓ |
| S63 | Kim et al. | 2008 | Retrospective | South Korea | Jan 2002- Dec 2005 | Cell culture, rapid antigen detection | Patients hospitalised for lower respiratory disease | ✓ | ✓ |  | ✓ |
| S64 | Shih et al. | 2015 | Prospective | Taiwan | Oct 2012-Jun 2013 | PCR, ESI-MS | Patients with acute RTIs treated at local outpatient clinic | ✓ | ✓ | ✓ | ✓ |

**Note:** 61 studies are included in Table S5. 3 studies (S07, S21, S43) were excluded after study IDs were assigned as they contained overlapping or modelled datasets (see methods for further information).<sup>7,13,42</sup>

#### A.6 Additional Results

**Figure S1:**
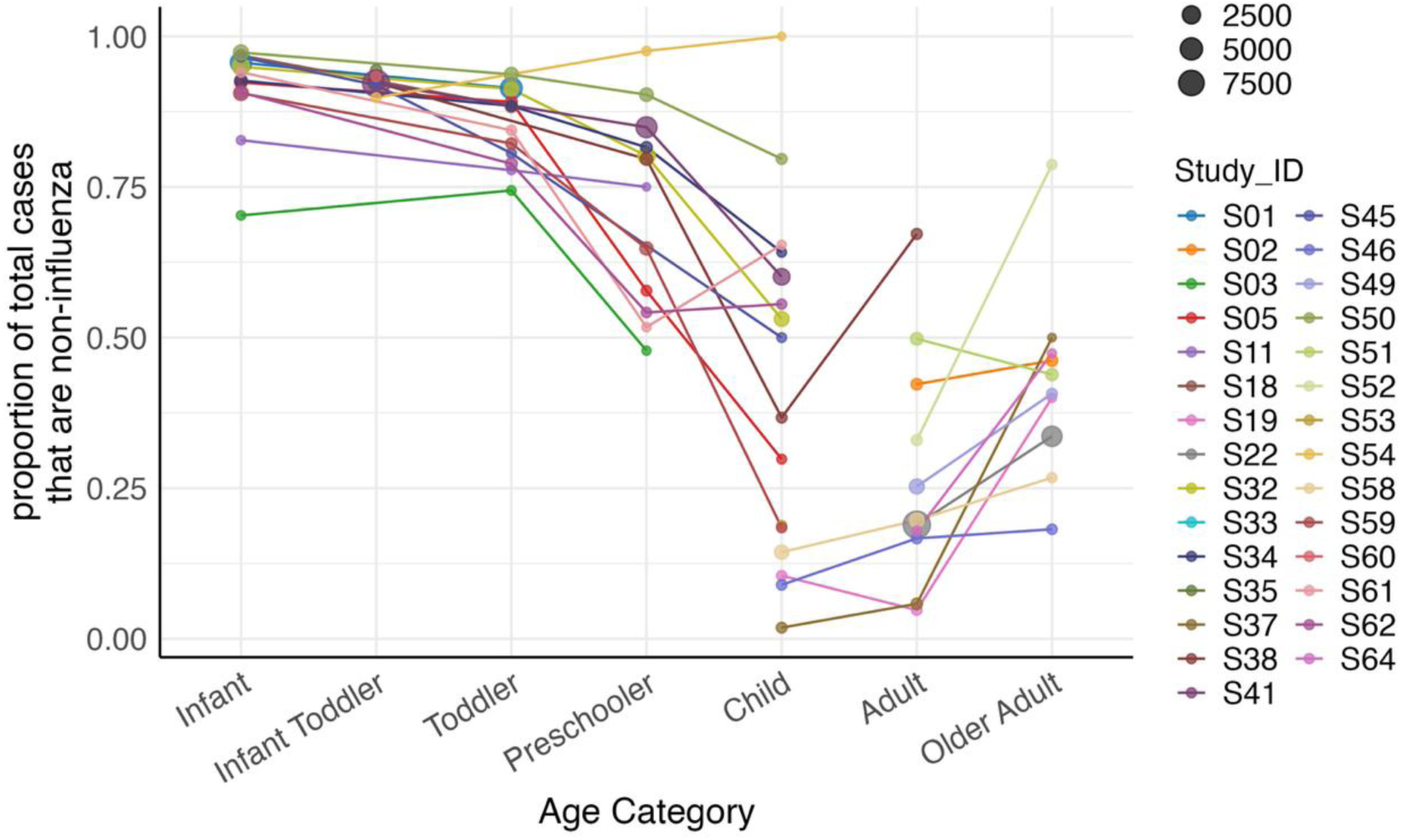
Proportion of detections that are non-influenza (PIV + HMPV + RSV), relative to total burden (PIV + HMPV + RSV + influenza). Each study is represented by a colour, where lines are shown to help visualise where datapoints are from the same study. The size of each datapoint represents the number of positive samples. Age groups: young infant (0–6 months); infant toddler (partial overlap across young infant and toddler groups); toddler (6–24 months); preschooler (2–5 years); child (5–18 years); adult (18–65 years); and older adult (65+ years).

**Figure S2:**
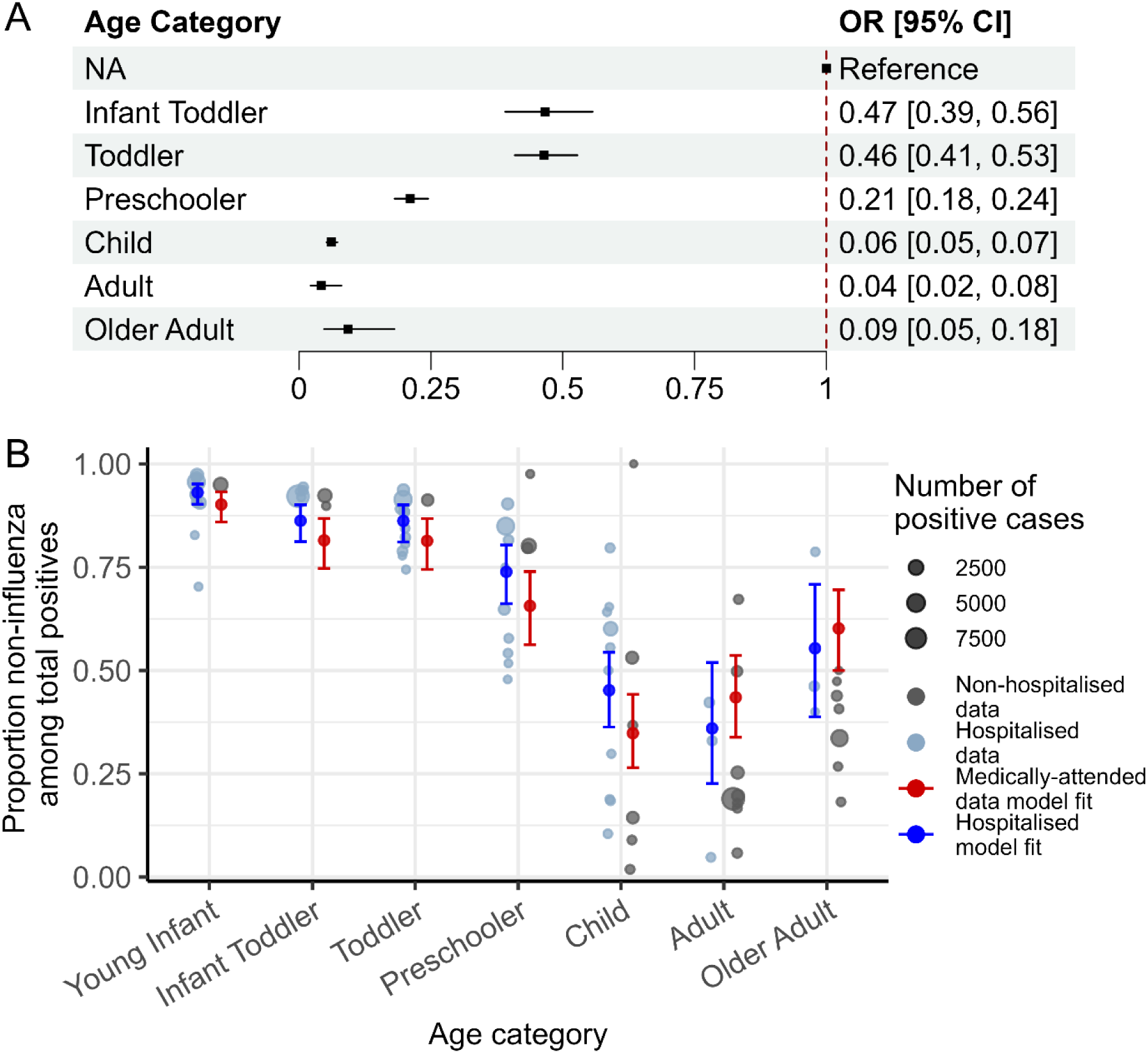
Age-specific hospitalised burden of non-influenza (HMPV, PIV and RSV), relative to influenza virus. A) Forest plot of model results showing odds ratios for the proportion of cases that are non-influenza for each age group, relative to the young infant reference group. B) Proportion of detections that are non-influenza (PIV + HMPV + RSV), relative to total burden (including influenza), comparing full dataset of medically-attended cases (hospitalised and non-hospitalised data; blue and grey points, model fit depicted in red) and hospitalised data subset (blue points only, model fit depicted in blue). Note that the hospitalised fit was produced by restricting to the 19 studies where the severity was classified as either hospitalised, ICU, or emergency department presentations (excluding emergency department attendances discharged without admission, but retaining those admitted or with unknown admission status). Age groups: young infant (0–6 months); infant toddler (partial overlap across young infant and toddler groups); toddler (6–24 months); preschooler (2–5 years); child (5–18 years); adult (18–65 years); and older adult (65+ years).

**Figure S3:**
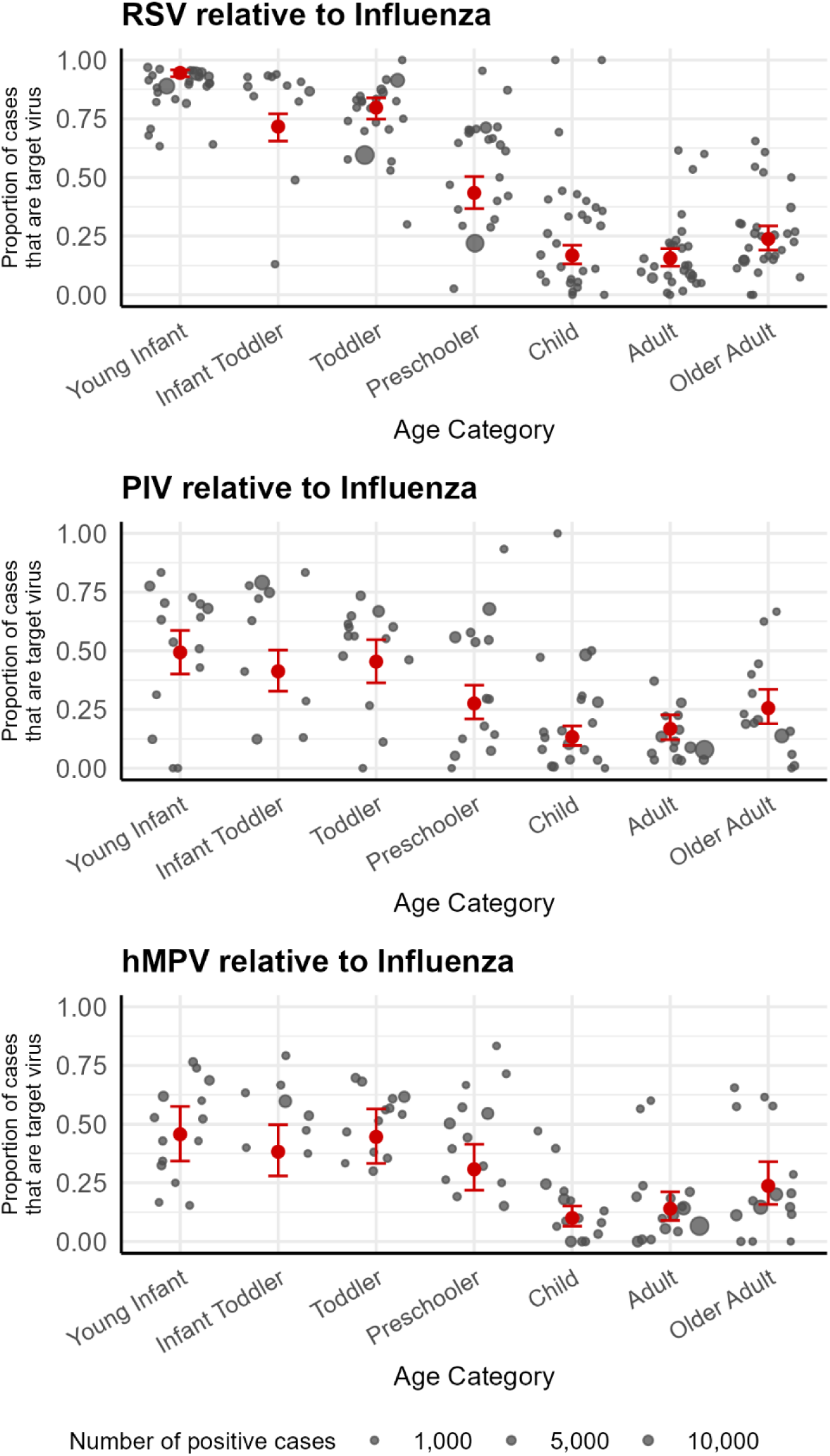
Age-specific burden of RSV, PIV and HMPV relative to influenza virus, with all reviewed studies (***n*** = 61). Absolute proportion of detections of each virus (RSV, HMPV, PIV), relative to the combined burden of each virus + influenza. Age groups: young infant (0-6 months); infant toddler (partial overlap across young infant and toddler groups); toddler (6-24 months); preschooler (2-5 years); child (5-18 years); adult (18-65 years); and older adult (65+ years).

**Figure S4:**
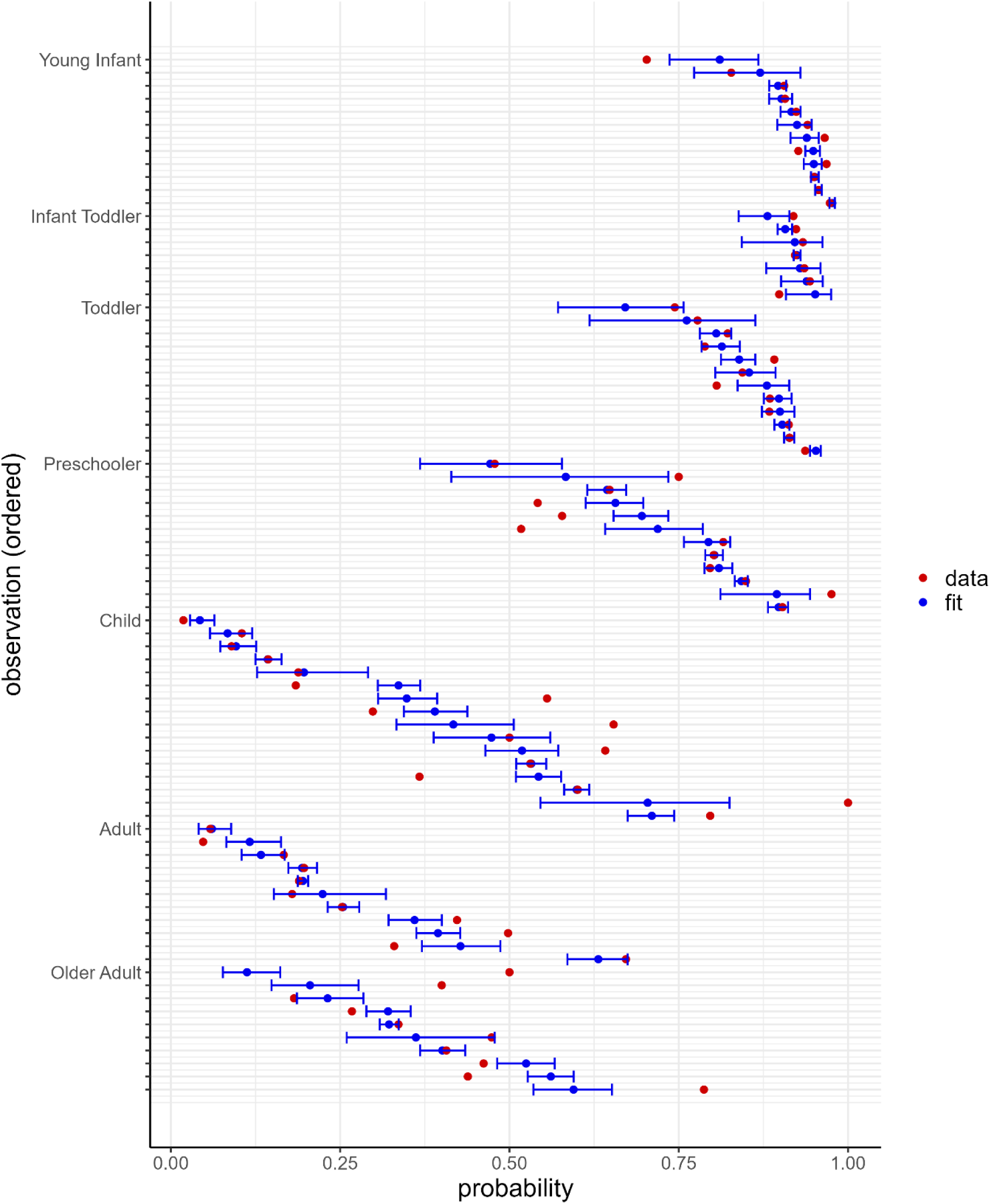
Illustration of model alignment with data for each observation, ordered by age group and study. Observed odds ratios for each study and age group, are shown as red points. For each observed point, we show the corresponding modelled estimate in blue (with 95% confidence interval), illustrating the degree of alignment between observed data and the fitted model. The model fit is shown for the dataset restricted to only studies that test for all four viruses of interest (influenza, hMPV, PIV and RSV), for any severity classification. Age groups: young infant (0–6 months); infant toddler (partial overlap across young infant and toddler groups); toddler (6–24 months); preschooler (2–5 years); child (5–18 years); adult (18–65 years); and older adult (65+ years).

### B Combination Vaccine Landscape Analysis

#### B.1 Methods

Using ClinicalTrials.gov and the World Health Organisation’s (WHO) International Clinical Trials Registry Platform (ICTRP), each virus (RSV, hMPV, PIV, influenza, and SARS-CoV-2) was searched individually under *condition/diseases*, and *“vaccine” OR “vaccination”* was used as the keyword under *intervention/treatment*. Clinical trials were included if they investigated vaccines designed to target two or more of the viruses of interest within the same vaccine and excluded if they focused only on a single pathogen (e.g. trivalent influenza vaccine). Two candidates targeting RSV and PIV-3 were excluded from the landscape snapshot as they represented older formulations (>15 years old).

For each eligible trial, we extracted the developer/sponsor, vaccine name/identifier, vaccine type (virus-like particles, nucleic acid, live-attenuated, subunit, inactivated or unknown), route of administration (intramuscular, intranasal, or unknown), intended age group, and targeted viral combination. A visual summary of the current combination vaccine landscape is provided in Figure S5.

#### B.2 Trial References

**Table S5.** Vaccine clinical trials included in the combination vaccine clinical trial landscape review. IM: intramuscular; IN: intranasal.

| Study Identifier | Developer | Name | Type | Trial Phase | Tested Cohort | Age Code | Year | Route | Link |
| --- | --- | --- | --- | --- | --- | --- | --- | --- | --- |
| **Influenza + SARS-CoV-2** |  |  |  |  |  |  |  |  |  |
| NCT05827926 | Moderna | mRNA-1083 | mRNA | 1, 2 | 18-79 years | A | 2023-2024 | IM | https://clinicaltrials.gov/study/NCT05827926 |
| NCT06097273 |  |  |  | 3 | 50+ years |  | 2023-2024 |  | https://clinicaltrials.gov/study/NCT06097273 |
| NCT06694389<br>JPRN-jRCT20312404<br>29 |  |  |  | 3 | 50+ years |  | 2024-2025 |  | https://clinicaltrials.gov/study/NCT06694389 |
| NCT06508320 |  |  |  | 2 | 50-65 years |  | 2024-2025 |  | https://clinicaltrials.gov/study/NCT06508320 |
| NCT06864143 |  |  |  | 2 | 18-65 years |  | 2025 |  | https://clinicaltrials.gov/study/NCT06864143 |
| NCT05375838 | Moderna | mRNA-1073 | mRNA | 1, 2 | 18-75 years | A | 2022 | IM | https://clinicaltrials.gov/study/NCT05375838 |
| NCT06178991<br>NCT05596734 | BioNTech / Pfizer | C5261002 | mRNA | 3 | 18-64 years | A | 2023-2024 | IM | https://clinicaltrials.gov/study/NCT06178991<br>https://clinicaltrials.gov/study/NCT05596734 |
| NCT05519839 | Novavax | CIC | Nanoparticle recombinant spike | 2 | 50-80 years | A | 2021-2022 | IM | https://clinicaltrials.gov/study/NCT04961541<br>https://clinicaltrials.gov/study/NCT05519839 |
| NCT06695130 | Sanofi Pasteur | VBT00002 | Recombinant | 1, 2 | 50+ years | A | 2024-2026 | IM | https://clinicaltrials.gov/study/NCT06695130 |
| NCT06695117 | Sanofi Pasteur | VBT00001 | Recombinant Inactivated | 1, 2 | 50+ years | A | 2024-2026 | IM | https://clinicaltrials.gov/study/NCT06695117 |
| ChiCTR2500104244 | West China Hospital of Sichuan University | Westvac C-19 + WSK-V102EF | Recombinant | 1 | 18+ years | A | 2025 | IM | https://trialsearch.who.int/Trial2.aspx?TrialID=ChiCTR2500104244 |

| hMPV + PIV-3 |  |  |  |  |  |  |  |  |  |
| --- | --- | --- | --- | --- | --- | --- | --- | --- | --- |
| NCT04144348 | Moderna | mRNA-16 53 | mRNA | 1b | 12-59 months, 18-49 years | A | 2023 | IM | https://clinicaltrials.gov/study/NCT04144348 |
| NCT03392389 |  |  | mRNA | 1 | 18-49 years |  | 2017-2019 | IM | https://clinicaltrials.gov/study/NCT03392389 |
| NCT06546423 | National Institute of Allergy and Infectious Diseases (NIAID) | B/HPIV3/HM PV-PreF-A / B/HPIV3/HM PV-F-B365 | Live-attenuated | 1 | 24-60 months | P | 2024-2025 | IN | https://clinicaltrials.gov/study/NCT06546423 |
| Influenza + RSV |  |  |  |  |  |  |  |  |  |
| NCT05585632 | Moderna | mRNA-1045 | mRNA | 1 | 50-75 years | A | 2022-2024 | IM | https://clinicaltrials.gov/study/NCT05585632 |
| RSV + hMPV |  |  |  |  |  |  |  |  |  |
| NCT05743881 | Moderna | mRNA-1365 | mRNA | 1 | 5-24 months | P | 2023-2026 | IM | https://clinicaltrials.gov/study/NCT05743881 |
| NCT05664334 | Icosavax | IVX-A12 | Virus-like particles | 1 | 60-75 years | A | 2022-2024 | IM | https://clinicaltrials.gov/study/NCT05664334 |
| NCT06481579 |  |  |  | 2 | 60+ years |  | 2024 | IM | https://clinicaltrials.gov/study/NCT06481579 |
| NCT05903183 |  |  |  | 2a | 60-85 years |  | 2023 | IM | https://clinicaltrials.gov/study/NCT05903183 |
| NCT06686654 | Sanofi Pasteur | VBD00009 | mRNA | 1, 2 | 60+ years | A | 2024-2027 | IM | https://clinicaltrials.gov/study/NCT06686654 |
| NCT06134648 | Sanofi Pasteur | VBD00003 | * | 1, 2a | 60+ years | A | 2023-2026 | IM | https://clinicaltrials.gov/study/NCT06134648 |
| NCT06237296 | Sanofi Pasteur | VAV00027 | mRNA | 1 | 18-49 years, 60+ years | A | 2024 | IM | https://clinicaltrials.gov/study/NCT06237296 |
| NCT06583031 | Sanofi Pasteur | VAV00039 | * | 1 | 60-75 years | A | 2024 | IM | https://clinicaltrials.gov/study/NCT06583031 |
| NCT06556147 | Vicebio Australia | VXB-241 | Subunit | 1 | 18-40 years, 60-83 years | A | 2024 | IM | https://clinicaltrials.gov/study/NCT06556147 |
| NCT06984094 | Clover Biopharmaceuticals | SCB-1022 | Recombinant | 1 | 60-85 Years | A | 2025-2026 | IM | https://clinicaltrials.gov/study/NCT06984094 |
| **Influenza + RSV + SARS-CoV-2** |  |  |  |  |  |  |  |  |  |
| NCT05585632 | Moderna | mRNA-1230 | mRNA | 1 | 50-75 years | A | 2022-2024 | IM | https://clinicaltrials.gov/study/NCT05585632 |
| **RSV + hMPV + PIV-3** |  |  |  |  |  |  |  |  |  |
| NCT06984094 | Clover Biopharmaceuticals | SCB-1033 | Recombinant | 1 | 60-85 years | A | 2024-2026 | IM | https://clinicaltrials.gov/study/NCT06984094 |
| NCT06604767 | Sanofi Pasteur | VBD00006 | * | 1 | 60+ years | A | 2024 | IM | https://clinicaltrials.gov/study/NCT06604767 |
| NCT06850051 | Sanofi Pasteur | VAV00030 | * | 1 | 18-49 years | A | 2025 | IM | https://clinicaltrials.gov/study/NCT06850051 |

\* Not stated in trial details

#### B.3 Visual Summary of the Viral Combination Vaccine Clinical Trial Landscape

**Figure S5:**
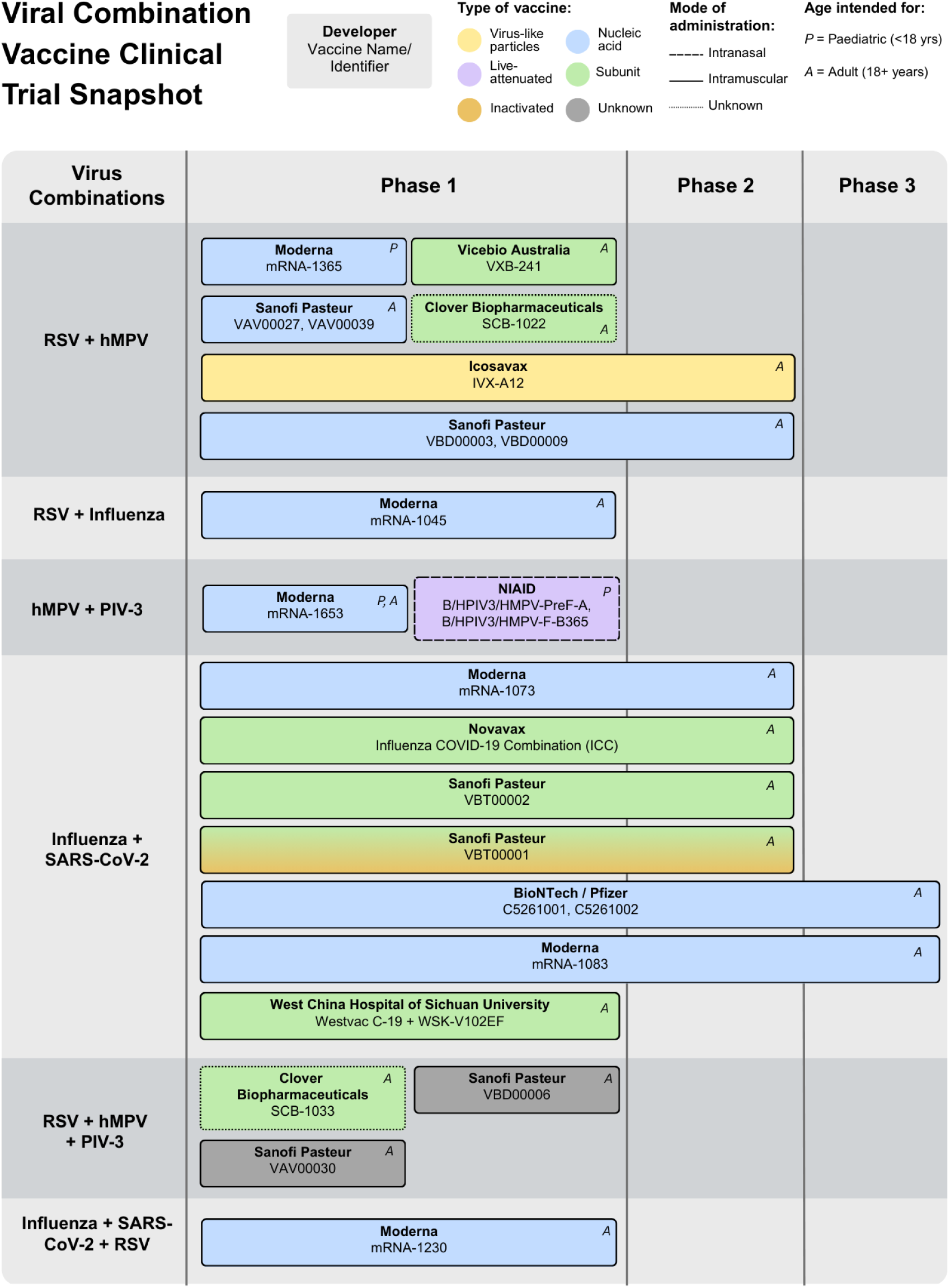
Viral combination vaccine clinical trial landscape analysis snapshot. Note: vaccine candidates from the same manufacturer at the same clinical trial phase are grouped together.

## References

1. World Health Organization. The top 10 causes of death. 2024. Accessed March 4, 2026. https://www.who.int/news-room/fact-sheets/detail/the-top-10-causes-of-death

2. Forum of International Respiratory Societies. The Global Impact of Respiratory Disease - 3rd *Edition*. European Respiratory Society; 2021. Accessed March 4, 2026. https://firsnet.org/wp-content/uploads/2025/01/FIRS_Master_09202021.pdf

3. Van Doorn HR, Yu H. Viral Respiratory Infections. In: Hunter’s Tropical Medicine and Emerging Infectious Diseases. Elsevier; 2020:284–288. doi:10.1016/B978-0-323-55512-8.00033-8

4. Agrawal S, Tran MT, Jennings TSK, et al. Changes in the innate immune response to SARS-CoV-2 with advancing age in humans. Immun Ageing. 2024;21(1):21. doi:10.1186/s12979-024-00426-3

5. Czaja CA, Miller L, Alden N, et al. Age-Related Differences in Hospitalization Rates, Clinical Presentation, and Outcomes Among Older Adults Hospitalized With Influenza—U.S. Influenza Hospitalization Surveillance Network (FluSurv-NET). Open Forum Infect Dis. 2019;6(7):ofz225. doi:10.1093/ofid/ofz225

6. Sarna M, Blyth CC, Taye BW, et al. Who is at risk of a respiratory syncytial virus hospitalisation? A linked, population-based birth cohort analysis in children aged less than 5 years. The Lancet Regional Health – Western Pacific. 2025;61. doi:10.1016/j.lanwpc.2025.101654

7. Kulkarni D, Cong B, Ranjini MJK, et al. The global burden of human metapneumovirus-associated acute respiratory infections in older adults: a systematic review and meta-analysis. The Lancet Healthy Longevity. 2025;6(2). doi:10.1016/j.lanhl.2024.100679

8. Farquharson KA, Anthony D, Menzies R, Homaira N. Burden of respiratory syncytial virus disease across the lifespan in Australia and New Zealand: a scoping review. Public Health. 2024;226:8–16. doi:10.1016/j.puhe.2023.10.031

9. Dobrzynski D, Branche AR, Falsey AR. Combination Seasonal Vaccines for Influenza, Respiratory Syncytial Virus, Severe Acute Respiratory Syndrome Coronavirus 2, and Other Pathogens. J Infect Dis. 2025;231(2):291–293. doi:10.1093/infdis/jiae507

10. Newall AT. The Value of Combination Vaccines in Children and Adults: More than the Sum of Their Parts. PharmacoEconomics. Published online February 17, 2026. doi:10.1007/s40273-026-01603-6

11. Income classifications for FY26 (July 1, 2025–June, 2026). The World Bank. Accessed February 24, 2026. https://datahelpdesk.worldbank.org/knowledgebase/articles/906519-world-bank-country-and-lending-groups

12. WebPlotDigitizer. Accessed February 24, 2026. https://automeris.io/

13. Bates D, Mächler M, Bolker B, Walker S. Fitting Linear Mixed-Effects Models Using lme4. Journal of Statistical Software. 2015;67:1–48. doi:10.18637/jss.v067.i01

14. Australian Bureau of Statistics. Q1 2024 Population Estimates (ERP), by State/Territory, Sex and Age [Data Explorer]. 2024. Accessed March 4, 2026. https://dataexplorer.abs.gov.au

15. CDC. Estimated US Flu Disease Burden. Flu Burden. March 19, 2026. Accessed April 14, 2026. https://www.cdc.gov/flu-burden/php/data-vis/index.html

16. Influenza | UKHSA data dashboard. Accessed April 14, 2026. https://ukhsa-dashboard.data.gov.uk/respiratory-viruses/influenza

17. Separation statistics by principal diagnosis (ICD-10-AM 12th edition), Australia, 2023-24. Australian Institute of Health and Welfare. July 8, 2025. Accessed February 25, 2026. https://www.aihw.gov.au/getmedia/8b58c4e0-7d13-4577-a905-5791fc784483/Principal-Diagnosis-cube-2023-24.xlsx

18. ClinicalTrials.gov. Accessed February 24, 2026. https://clinicaltrials.gov/

19. International Clinical Trials Registry Platform (ICTRP). Accessed February 24, 2026. https://trialsearch.who.int/

20. Aminisani N, Wood T, Jelley L, Wong C, Sue Huang Q. The Burden of Human Metapneumovirus- and Influenza-Associated Hospitalizations in Adults in New Zealand, 2012–2015. The Journal of Infectious Diseases. 2024;230(4):933–943. doi:10.1093/infdis/jiae064

21. Prasad N, Trenholme AA, Huang QS, Duque J, Grant CC, Newbern EC. Respiratory Virus-related Emergency Department Visits and Hospitalizations Among Infants in New Zealand. Pediatric Infectious Disease Journal. 2020;39(8):e176–e182. doi:10.1097/INF.0000000000002681

22. Aguilar Figueroa D, Pérez-Gimeno G, Núñez O, Monge S, The SiVIRA Surveillance and Vaccine Effectiveness Working Group. Burden of Severe Disease Associated With Influenza, SARS-CoV-2 and RSV in Spain During the 2024–2025 Winter Season. Influenza Resp Viruses. 2025;19(11):e70190. doi:10.1111/irv.70190

23. Kenmoe S, Nair H. The disease burden of respiratory syncytial virus in older adults. Curr Opin Infect Dis. 2024;37(2):129–136. doi:10.1097/QCO.0000000000001000

24. Langer J, Welch VL, Moran MM, et al. High Clinical Burden of Influenza Disease in Adults Aged ≥ 65 Years: Can We Do Better? A Systematic Literature Review. Adv Ther. 2023;40(4):1601–1627. doi:10.1007/s12325-023-02432-1

25. Sobanjo-ter Meulen A, Gutierrez AV, Ruiz O, Eeuwijk J, Vroling H, Kanesa-thasan N. The Burden of Human Metapneumovirus (hMPV) Disease in Older and High-Risk Adults in Developed Countries: A Systematic Literature Review. Infect Dis Ther. 2025;14(8):1917–1933. doi:10.1007/s40121-025-01187-2

26. Wang X, Li Y, Deloria-Knoll M, et al. Global burden of acute lower respiratory infection associated with human metapneumovirus in children under 5 years in 2018: a systematic review and modelling study. The Lancet Global Health. 2021;9(1):e33–e43. doi:10.1016/S2214-109X(20)30393-4

27. Martyn O, Vlasich C, Khariv V, Boudewyn LC, Openshaw PJM, Kramer R. Global Epidemiology and Disease Burden of Human Parainfluenza Virus in Adults: A Systematic Review. Rev Med Virol. 2026;36(1):e70105. doi:10.1002/rmv.70105

28. Trusinska D, Lee B, Ferdous S, et al. Real-world evidence on RSV vaccine uptake, effectiveness, and safety in older adults: a systematic review and meta-analysis. Lancet Reg Health Eur. 2026;64:101623. doi:10.1016/j.lanepe.2026.101623

29. Lagler FB, Hirschfeld S, Kindblom JM. Challenges in clinical trials for children and young people. Published online April 1, 2021. doi:10.1136/archdischild-2019-318676

30. Medicine TLR. COVID-19 pandemic disturbs respiratory virus dynamics. The Lancet Respiratory Medicine. 2022;10(8):725. doi:10.1016/S2213-2600(22)00255-7

31. Plotkin SA. Why Combination Vaccines Are Necessary. The Pediatric Infectious Disease Journal. 2024;43(11):1046. doi:10.1097/INF.0000000000004476

## References

S01 Sarna M, Le H, Taye BW, et al. Clinical outcomes and severity of laboratory-confirmed RSV compared with influenza, parainfluenza and human metapneumovirus in Australian children attending secondary care. BMJ Open Resp Res. 2024;11(1):e002613. doi:10.1136/bmjresp-2024-002613

S02 Minney-Smith CA, Selvey LA, Levy A, Smith DW. Post-pandemic influenza A/H1N1pdm09 is associated with more severe outcomes than A/H3N2 and other respiratory viruses in adult hospitalisations. Epidemiol Infect. 2019;147:e310. doi:10.1017/S095026881900195X

S03 Amer HM, Alshaman MS, Farrag MA, Hamad ME, Alsaadi MM, Almajhdi FN. Epidemiology of 11 respiratory RNA viruses in a cohort of hospitalized children in Riyadh, Saudi Arabia. Journal of Medical Virology. 2016;88(6):1086–1091. doi:10.1002/jmv.24435

S04 Widmer K, Zhu Y, Williams JV, Griffin MR, Edwards KM, Talbot HK. Rates of Hospitalizations for Respiratory Syncytial Virus, Human Metapneumovirus, and Influenza Virus in Older Adults. The Journal of Infectious Diseases. 2012;206(1):56–62. doi:10.1093/infdis/jis309

S05 Choi E, Ha KS, Song DJ, Lee JH, Lee KC. Clinical and laboratory profiles of hospitalized children with acute respiratory virus infection. Korean J Pediatr. 2018;61(6):180. doi:10.3345/kjp.2018.61.6.180

S06 Shinkai M, Ota S, Ishikawa N, et al. Burden of respiratory syncytial virus, human metapneumovirus and influenza virus infections in Japanese adults in the Hospitalized Acute Respiratory Tract Infection study. Respiratory Investigation. 2024;62(4):717–725. doi:10.1016/j.resinv.2024.05.015

S07 Aminisani N, Wood T, Jelley L, Wong C, Sue Huang Q. The Burden of Human Metapneumovirus- and Influenza-Associated Hospitalizations in Adults in New Zealand, 2012–2015. The Journal of Infectious Diseases. 2024;230(4):933–943. doi:10.1093/infdis/jiae064

S08 Nolen LD, Seeman S, Desnoyers C, et al. Respiratory syncytial virus and influenza hospitalizations in Alaska native adults. Journal of Clinical Virology. 2020;127:104347. doi:10.1016/j.jcv.2020.104347

S09 Wolf DG, Greenberg D, Kalkstein D, et al. Comparison of Human Metapneumovirus, Respiratory Syncytial Virus and Influenza A Virus Lower Respiratory Tract Infections in Hospitalized Young Children: The Pediatric Infectious Disease Journal. 2006;25(4):320–324. doi:10.1097/01.inf.0000207395.80657.cf

S10 Sominina A, Danilenko D, Komissarov A, et al. Age-Specific Etiology of Severe Acute Respiratory Infections and Influenza Vaccine Effectivity in Prevention of Hospitalization in Russia, 2018–2019 Season. J Epidemiol Glob Health. 2021;11(4):413–425. doi:10.1007/s44197-021-00009-1

S11 Jennings LC, Anderson TP, Werno AM, Beynon KA, Murdoch DR. Viral Etiology of Acute Respiratory Tract Infections in Children Presenting to Hospital: Role of Polymerase Chain Reaction and Demonstration of Multiple Infections. The Pediatric Infectious Disease Journal. 2004;23(11):1003–1007. doi:10.1097/01.inf.0000143648.04673.6c

S12 Arashiro T, Kramer R, Jin J, Kano M, Wang F, Miyairi I. Inpatient Burden of Respiratory Syncytial Virus Infection and Influenza in Children Younger Than 5 Years in Japan, 2011–2022: A Database Study. Influenza Resp Viruses. 2024;18(11):e70045. doi:10.1111/irv.70045

S13 Davis W, Duque J, Huang QS, et al. Sensitivity and specificity of surveillance case definitions in detection of influenza and respiratory syncytial virus among hospitalized patients, New Zealand, 2012–2016. Journal of Infection. 2022;84(2):216–226. doi:10.1016/j.jinf.2021.12.012

S14 Symes R, Keddie SH, Walker J, et al. Burden of respiratory syncytial virus infection in older adults hospitalised in England during 2023/24. Journal of Infection. 2025;91(3):106570. doi:10.1016/j.jinf.2025.106570

S15 Ajayi-Obe EK, Coen PG, Handa R, et al. Influenza A and respiratory syncytial virus hospital burden in young children in East London. Epidemiol Infect. 2008;136(8):1046–1058. doi:10.1017/S0950268807009557

S16 Buchan SA, Chung H, Karnauchow T, et al. Characteristics and Outcomes of Young Children Hospitalized With Laboratory-confirmed Influenza or Respiratory Syncytial Virus in Ontario, Canada, 2009–2014. Pediatric Infectious Disease Journal. 2019;38(4):362–369. doi:10.1097/INF.0000000000002164

S17 Chorazka M, Flury D, Herzog K, Albrich WC, Vuichard-Gysin D. Clinical outcomes of adults hospitalized for laboratory confirmed respiratory syncytial virus or influenza virus infection. Chen TH, ed. PLoS ONE. 2021;16(7):e0253161. doi:10.1371/journal.pone.0253161

S18 Trenholme AA, Best EJ, Vogel AM, Stewart JM, Miller CJ, Lennon DR. Respiratory virus detection during hospitalisation for lower respiratory tract infection in children under 2 years in South Auckland, New Zealand. J Paediatrics Child Health. 2017;53(6):551–555. doi:10.1111/jpc.13529

S19 Chan PA, Mermel LA, Andrea SB, et al. Distinguishing Characteristics between Pandemic 2009–2010 Influenza A (H1N1) and Other Viruses in Patients Hospitalized with Respiratory Illness. Pekosz A, ed. PLoS ONE. 2011;6(9):e24734. doi:10.1371/journal.pone.0024734

S20 Ackerson B, Tseng HF, Sy LS, et al. Severe Morbidity and Mortality Associated With Respiratory Syncytial Virus Versus Influenza Infection in Hospitalized Older Adults. Clinical Infectious Diseases. 2019;69(2):197–203. doi:10.1093/cid/ciy991

S22 Al-Romaihi HE, Smatti MK, Ganesan N, et al. Epidemiology of respiratory infections among adults in Qatar (2012-2017). Dijkman R, ed. PLoS ONE. 2019;14(6):e0218097. doi:10.1371/journal.pone.0218097

S23 Al-Shehri MA, Sadeq A, Quli K. Bronchiolitis in Abha, Southwest Saudi Arabia: viral etiology and predictors for hospital admission. West African Journal of Medicine. 2006;24(4):299–304. doi:10.4314/wajm.v24i4.28193

S24 Almehmadi MM, Alharbi FA, Shawush AK, et al. Molecular identification of respiratory syncytial virus, seasonal influenza viruses, and SARS-CoV-2 in respiratory tract infections: A cross-sectional study in Taif city. SMJ. 2025;46(10):1202–1208. doi:10.15537/smj.2025.46.10.20250133

S25 Aminisani N, Fanslow B, Wood T, et al. The Burden of HMPV- and Influenza-Associated Hospitalizations in Adults in New Zealand Before and After the COVID-19 Pandemic, 2012–2023. The Journal of Infectious Diseases. 2025;232(Supplement_1):S47–S58. doi:10.1093/infdis/jiaf150

S26 Balas W, Balas K, Gancarczyk M, et al. Clinical Characteristics of Paediatric RSV, Influenza, and SARS-CoV-2 Infections: Insights from Three Consecutive Seasons. Viruses. 2025;17(11):1403. doi:10.3390/v17111403

S27 Begley KM, Leis AM, Petrie JG, et al. Epidemiology of Respiratory Syncytial Virus in Adults and Children With Medically Attended Acute Respiratory Illness Over Three Seasons. Clinical Infectious Diseases. 2024;79(4):1039–1045. doi:10.1093/cid/ciae303

S28 Begley KM, Monto AS, Lamerato LE, et al. Prevalence and Clinical Outcomes of Respiratory Syncytial Virus vs Influenza in Adults Hospitalized With Acute Respiratory Illness From a Prospective Multicenter Study. Clinical Infectious Diseases. 2023;76(11):1980–1988. doi:10.1093/cid/ciad031

S29 Bøås H, Seppälä E, Veneti L, et al. Changed epidemiology of influenza and RSV hospitalizations after the emergence of SARS-CoV-2 in Norway, 2017–2024. Annals of Epidemiology. 2025;108:77–84. doi:10.1016/j.annepidem.2025.06.010

S30 Bosis S, Esposito S, Niesters HGM, et al. Role of respiratory pathogens in infants hospitalized for a first episode of wheezing and their impact on recurrences. Clinical Microbiology and Infection. 2008;14(7):677–684. doi:10.1111/j.1469-0691.2008.02016.x

S31 Bourgeois FT, Valim C, McAdam AJ, Mandl KD. Relative Impact of Influenza and Respiratory Syncytial Virus in Young Children. Pediatrics. 2009;124(6):e1072–e1080. doi:10.1542/peds.2008-3074

S32 Burrell R, Saravanos GL, Kesson A, et al. Respiratory virus detections in children presenting to an Australian paediatric referral hospital pre-COVID-19 pandemic, January 2014 to December 2019. Lau EH, ed. PLoS ONE. 2025;20(1):e0313504. doi:10.1371/journal.pone.0313504

S33 De Luca M, D’Amore C, Romani L, et al. Severe viral respiratory infections in the pre-COVID era: A 5-year experience in two pediatric intensive care units in Italy. Influenza Resp Viruses. 2023;17(1):e13038. doi:10.1111/irv.13038

S34 Fagbo SF, Garbati MA, Hasan R, et al. Acute viral respiratory infections among children in MERS-endemic Riyadh, Saudi Arabia, 2012–2013. Journal of Medical Virology. 2017;89(2):195–201. doi:10.1002/jmv.24632

S35 García-García ML, Calvo C, Pozo F, Villadangos PA, Pérez-Breña P, Casas I. Spectrum of Respiratory Viruses in Children With Community-acquired Pneumonia. Pediatric Infectious Disease Journal. 2012;31(8):808–813. doi:10.1097/INF.0b013e3182568c67

S36 Gooskens J, Van Der Ploeg V, Sukhai RN, Vossen AC, Claas EC, Kroes AC. Clinical evaluation of viral acute respiratory tract infections in children presenting to the emergency department of a tertiary referral hospital in the Netherlands. BMC Pediatr. 2014;14(1):297. doi:10.1186/s12887-014-0297-0

S37 Hombrouck A, Sabbe M, Casteren V, et al. Viral aetiology of influenza-like illness in Belgium during the influenza A(H1N1)2009 pandemic. Eur J Clin Microbiol Infect Dis. 2012;31(6):999–1007. doi:10.1007/s10096-011-1398-4

S38 Kim JK. Epidemiology of Respiratory Viral Infection Using Multiplex RT-PCR in Cheonan, Korea (2006-2010). J Microbiol Biotechnol. 2013;23(2):267–273. doi:10.4014/jmb.1212.12050

S39 Kusel MMH, De Klerk NH, Holt PG, Kebadze T, Johnston SL, Sly PD. Role of Respiratory Viruses in Acute Upper and Lower Respiratory Tract Illness in the First Year of Life: A Birth Cohort Study. The Pediatric Infectious Disease Journal. 2006;25(8):680–686. doi:10.1097/01.inf.0000226912.88900.a3

S40 Lauderdale TL, Chang FY, Ben RJ, et al. Etiology of community acquired pneumonia among adult patients requiring hospitalization in Taiwan. Respiratory Medicine. 2005;99(9):1079–1086. doi:10.1016/j.rmed.2005.02.026

S41 Pneumonia and Respiratory Disease Study Group of Korean Academy of Pediatric Allergy and Respiratory Disease, Lee E, Kim CH, et al. Annual and seasonal patterns in etiologies of pediatric community-acquired pneumonia due to respiratory viruses and Mycoplasma pneumoniae requiring hospitalization in South Korea. BMC Infect Dis. 2020;20(1):132. doi:10.1186/s12879-020-4810-9

S42 Mansbach JM, McAdam AJ, Clark S, et al. Prospective Multicenter Study of the Viral Etiology of Bronchiolitis in the Emergency Department. Academic Emergency Medicine. 2008;15(2):111–118. doi:10.1111/j.1553-2712.2007.00034.x

S43 Prasad N, Trenholme AA, Huang QS, Duque J, Grant CC, Newbern EC. Respiratory Virus-related Emergency Department Visits and Hospitalizations Among Infants in New Zealand. Pediatric Infectious Disease Journal. 2020;39(8):e176–e182. doi:10.1097/INF.0000000000002681

S44 Puzelli S, Valdarchi C, Ciotti M, et al. Viral causes of influenza-like illness: Insight from a study during the winters 2004–2007. Journal of Medical Virology. 2009;81(12):2066–2071. doi:10.1002/jmv.21610

S45 Richter J, Panayiotou C, Tryfonos C, et al. Aetiology of Acute Respiratory Tract Infections in Hospitalised Children in Cyprus. Sugrue RJ, ed. PLoS ONE. 2016;11(1):e0147041. doi:10.1371/journal.pone.0147041

S46 Russell KE, Fowlkes A, Stockwell MS, et al. Comparison of outpatient medically attended and community-level influenza-like illness—New York City, 2013-2015. Influenza Resp Viruses. 2018;12(3):336–343. doi:10.1111/irv.12540

S47 Tramuto F, Maida CM, Napoli G, et al. Burden and viral aetiology of influenza-like illness and acute respiratory infection in intensive care units. Microbes and Infection. 2016;18(4):270–276. doi:10.1016/j.micinf.2015.11.008

S48 Turunen R, Koistinen A, Vuorinen T, et al. The first wheezing episode: respiratory virus etiology, atopic characteristics, and illness severity. Pediatric Allergy Immunology. 2014;25(8):796–803. doi:10.1111/pai.12318

S49 Varghese BM, Dent E, Chilver M, Cameron S, Stocks NP. Epidemiology of viral respiratory infections in Australian working-age adults (20–64 years): 2010–2013. Epidemiol Infect. 2018;146(5):619–626. doi:10.1017/S0950268818000286

S50 Vila J, Lera E, Andrés C, et al. The burden of non-SARS-CoV2 viral lower respiratory tract infections in hospitalized children in Barcelona (Spain): A long-term, clinical, epidemiologic and economic study. Influenza Resp Viruses. 2023;17(1):e13085. doi:10.1111/irv.13085

S51 Visseaux B, Burdet C, Voiriot G, et al. Prevalence of respiratory viruses among adults, by season, age, respiratory tract region and type of medical unit in Paris, France, from 2011 to 2016. Schanzer DL, ed. PLoS ONE. 2017;12(7):e0180888. doi:10.1371/journal.pone.0180888

S52 Walker E, Ison MG. Respiratory viral infections among hospitalized adults: experience of a single tertiary healthcare hospital. Influenza Resp Viruses. 2014;8(3):282–292. doi:10.1111/irv.12237

S53 Windsor WJ, Lamb MM, Dominguez SR, Mistry RD, Rao S. Clinical characteristics and illness course based on pathogen among children with respiratory illness presenting to an emergency department. Journal of Medical Virology. 2022;94(12):6103–6110. doi:10.1002/jmv.28031

S54 Yen CY, Wu WT, Chang CY, et al. Viral etiologies of acute respiratory tract infections among hospitalized children – A comparison between single and multiple viral infections. *Journal of Microbiology*, Immunology and Infection. 2019;52(6):902–910. doi:10.1016/j.jmii.2019.08.013

S55 Zimmerman RK, Rinaldo CR, Nowalk MP, et al. Viral infections in outpatients with medically attended acute respiratory illness during the 2012–2013 influenza season. BMC Infect Dis. 2015;15(1):87. doi:10.1186/s12879-015-0806-2

S56 Zimmerman RK, Rinaldo CR, Nowalk MP, et al. Influenza and other respiratory virus infections in outpatients with medically attended acute respiratory infection during the 2011-12 influenza season. Influenza Resp Viruses. 2014;8(4):397–405. doi:10.1111/irv.12247

S57 Loubet P, Guitton S, Rolland S, et al. Characteristics of Human Metapneumovirus Infection Compared to Respiratory Syncytial Virus and Influenza Infections in Adults Hospitalized for Influenza-Like Illness in France, 2012–2022. The Journal of Infectious Diseases. 2025;232(Supplement_1):S93–S100. doi:10.1093/infdis/jiaf082

S58 Antón A, Marcos MA, Torner N, et al. Virological surveillance of influenza and other respiratory viruses during six consecutive seasons from 2006 to 2012 in Catalonia, Spain. Clinical Microbiology and Infection. 2016;22(6):564.e1-564.e9. doi:10.1016/j.cmi.2016.02.007

S59 Bosch Castells V, Mira-Iglesias A, López-Labrador FX, et al. Pediatric Respiratory Hospitalizations in the Pre-COVID-19 Era: The Contribution of Viral Pathogens and Comorbidities to Clinical Outcomes, Valencia, Spain. Viruses. 2024;16(10):1519. doi:10.3390/v16101519

S60 Chen YW, Huang YC, Ho TH, Huang CG, Tsao KC, Lin TY. Viral etiology of bronchiolitis among pediatric inpatients in northern Taiwan with emphasis on newly identified respiratory viruses. *Journal of Microbiology*, Immunology and Infection. 2014;47(2):116–121. doi:10.1016/j.jmii.2012.08.012

S61 Cohen S, Dabaja-Younis H, Etshtein L, et al. Burden of viral respiratory infections in the pediatric intensive care unit: age, virus distribution, and the impact of the COVID-19 pandemic. Eur J Pediatr. 2024;184(1):88. doi:10.1007/s00431-024-05914-8

S62 De Conto F, Conversano F, Medici MC, et al. Epidemiology of human respiratory viruses in children with acute respiratory tract infection in a 3-year hospital-based survey in Northern Italy. Diagnostic Microbiology and Infectious Disease. 2019;94(3):260–267. doi:10.1016/j.diagmicrobio.2019.01.008

S63 Kim YK, Nyambat B, Hong YS, Lee CG, Lee JW, Kilgore PE. Burden of viral respiratory disease hospitalizations among children in a community of Seoul, Republic of Korea, 1995 – 2005. Scandinavian Journal of Infectious Diseases. 2008;40(11-12):946–953. doi:10.1080/00365540802398937

S64 Shih HI, Wang HC, Su IJ, et al. Viral Respiratory Tract Infections in Adult Patients Attending Outpatient and Emergency Departments, Taiwan, 2012–2013: A PCR/Electrospray Ionization Mass Spectrometry Study. Medicine. 2015;94(38):e1545. doi:10.1097/MD.0000000000001545

64. Australian Bureau of Statistics. Q1 2024 Population Estimates (ERP), by State/Territory, Sex and Age [Data Explorer]. 2024. Accessed March 4, 2026. https://dataexplorer.abs.gov.au/vis?tm=quarterly%20population&pg=0&df[ds]=ABS_ABS_TOPICS&df[id]=ERP_Q&df[ag]=ABS&df[vs]=1.0.0&hc[Frequency]=Quarterly&pd=,&dq=….Q&to[TIME_PERIOD]=false&vw=tb

65. CDC. Estimated US Flu Disease Burden. Flu Burden. March 19, 2026. Accessed April 14, 2026. https://www.cdc.gov/flu-burden/php/data-vis/index.html

66. Influenza | UKHSA data dashboard. Accessed April 14, 2026. https://ukhsa-dashboard.data.gov.uk/respiratory-viruses/influenza

67. International Classifications in Australia. Australian Institute of Health and Welfare. October 23, 2024. Accessed March 4, 2026. https://www.aihw.gov.au/about-us/international-collaboration/australian-collaborating-centre-for-who/international-classifications-in-australia

68. Nazareno AL, Muscatello DJ, Turner RM, Wood JG, Moore HC, Newall AT. Modelled estimates of hospitalisations attributable to respiratory syncytial virus and influenza in Australia, 2009–2017. Influenza and Other Respiratory Viruses. 2022;16(6):1082–1090. doi:10.1111/irv.13003

69. Separation statistics by principal diagnosis (ICD-10-AM 12th edition), Australia, 2023-24. Australian Institute of Health and Welfare. July 8, 2025. Accessed February 25, 2026. https://www.aihw.gov.au/getmedia/8b58c4e0-7d13-4577-a905-5791fc784483/Principal-Diagnosis-cube-2023-24.xlsx

